# Combined epidemiological and genomic analysis of nosocomial SARS-CoV-2 transmission identifies community social distancing as the dominant intervention reducing outbreaks

**DOI:** 10.1101/2020.11.17.20232827

**Authors:** Luke B Snell, Chloe L Fisher, Usman Taj, Blair Merrick, Adela Alcolea-Medina, Themoula Charalampous, Adrian W Signell, Harry D Wilson, Gilberto Betancor, Mark Tan Kia Ik, Emma Cunningham, Penelope R Cliff, Suzanne Pickering, Rui Pedro Galao, Rahul Batra, Stuart J D Neil, Michael H Malim, Katie J Doores, Sam T Douthwaite, Gaia Nebbia, Jonathan D Edgeworth, Ali R Awan, The COVID-19 Genomics UK (COG-UK) consortium

## Abstract

Many healthcare facilities report SARS-CoV-2 outbreaks but transmission analysis is complicated by the high prevalence of infection and limited viral genetic diversity. The contribution of different vectors to nosocomial infection or the effectiveness of interventions is therefore currently unclear. Detailed epidemiological and viral nanopore sequence data were analysed from 574 consecutive patients with a PCR positive SARS-CoV-2 test between March 13th and March 31st, when the pandemic first impacted on a large, multisite healthcare institution in London. During this time the first major preventative interventions were introduced, including progressive community social distancing (CSD) policies leading to mandatory national lockdown, exclusion of hospital visitors, and introduction of universal surgical facemask-use by healthcare-workers (HCW). Incidence of nosocomial cases, community SARS-CoV-2 cases and infection in a cohort of 228 HCWs followed the same dynamic course, decreasing shortly after introduction of CSD measures and prior to the main hospital-based interventions. We investigated clusters involving nosocomial cases based on overlapping ward-stays during the 14-day incubation period and SARS-CoV-2 genome sequence similarity. Our method placed 63 (79%) of 80 sequenced probable and definite nosocomial cases into 14 clusters containing a median of 4 patients (min 2, max 19) No genetic support was found for the majority of epidemiological clusters (31/44, 70%) and genomics revealed multiple contemporaneous outbreaks within single epidemiological clusters. We included a measure of hospital enrichment compared to community cases to increase confidence in our clusters, which were 1-14 fold enriched. Applying genomics, we could provide a robust estimate of the incubation period for nosocomial transmission, with a median lower bound and upper bound of 6 and 9 days respectively. Six (43%) clusters spanned multiple wards, with evidence of cryptic transmission, and community-onset cases could not be identified in more than half the clusters, particularly on the elective hospital site, implicating HCW as vectors of transmission. Taken together these findings suggest that CSD had the dominant impact on reducing nosocomial transmission by reducing HCW infection.

## INTRODUCTION

Severe acute respiratory syndrome coronavirus 2 (SARS-CoV-2) was first reported in Wuhan, China in December 2019 (1), since when over 40 million infections and 1 million deaths have been reported worldwide (2). New cases in the UK peaked during the first wave after implementation of progressive community social distancing (CSD) (3), starting with an announcement on March 16th that, with few exceptions, the whole UK population must stay at home, later being made mandatory on March 23rd. COVID-19 hospital admission then peaked around 8 days later (4).

The first few weeks of the pandemic were unique, due to introduction of SARS-CoV-2 into populations without prior immunity. Society and healthcare organisations were adapting to the pandemic for the first time with frequent new infection control policies and behavioural changes while incidence was still accelerating. During the first wave, elective hospital admissions were often postponed (5). Currently, hospitals around the world either have, or are preparing for, a further influx of COVID-19 patients, while restarting and rescheduling postponed non-COVID-19 work. It is therefore important to identify evidence from these first few weeks to help healthcare organisations efficiently plan for any subsequent increase in cases.

Most COVID-19 transmission studies to date have utilised epidemiological analysis alone to identify outbreaks, including those from the UK (6) (7), China (8), France (9) and South Korea (10). The main limitation with using epidemiology alone is that when point prevalence is high, for instance at 2.2% in London during the first wave in April 2020 (11), this increases the chance two people in epidemiological contact are independent cases. Furthermore, a wide incubation period of between 2 and 14 days (12,13) makes it difficult to define hospital-onset cases as hospital-acquired, as even when they arise several days after admission to hospital the transmission event may still have occurred in the community.

To improve the confidence in detecting hospital clusters, epidemiological analysis can be supplemented with genomic data obtained by sequencing viral isolates. For COVID-19, thus far only two published studies have used genomic sequencing to analyse hospital transmission (14) (15). However, application of genomic data to the spread of SARS-CoV-2 is not straightforward, particularly as during early stages of the pandemic, genetic diversity was low with less than 200 mutations registered in the international GISAID database by April 2020 (16). SARS-CoV-2 also has a low mutation rate with only 2-3 mutations per genome per month (17). These factors increase the chance that two people infected with virus sharing identical genetic sequence are not closely epidemiologically linked cases. However, epidemiological linkage combined with viral sequence data can provide stronger evidence for a COVID-19 transmission cluster than would be possible with either source of data alone.

Nosocomial infection is reported to account for around 10-20% of all confirmed cases (18) (19) (6) (20) often in designated outbreaks (6) (7) (8) (9) (10) and with a crude death rate of up to 30% (19). The most likely routes of transmission are between patients and involving healthcare workers (HCW), with potential for all combinations of directionality to occur within the same nosocomial cluster. In each case, the transmitting individual may be either symptomatic or asymptomatic. There is also potential for super-spreading events (21). It is important to determine the contribution of each pathway to help prioritise resourcing of interventions. Most hospitals excluded visitors, who are an alternate source for introducing infection, and although the environment can become contaminated with infectious droplets, cleaning policies effectively removes virus from the environment (22), so fomite spread remains an unproven mode of transmission (23). Airborne transmission via aerosols remains controversial (23).

The aim of this study was to develop a methodology for combining epidemiological and genomics analysis of SARS-CoV-2 transmission to help determine the pathways for nosocomial transmission during the first wave of the pandemic. During this time there was a high incidence in the community (11) and high incidence of COVID-19 infection amongst healthcare workers (HCWs) as judged by subsequent seroconversion (24) (25).

## RESULTS

### Setting, clinical characteristics and epidemiology of cases in the first wave of the pandemic

SARS-CoV-2 PCR testing for all hospital admissions and inpatients with compatible clinical symptoms of new fever or cough was commenced on March 13th in line with national UK recommendations. Between 80 and 150 SARS-CoV-2 PCR tests were performed per day until the end of April when testing increased further (Table S1), with 637 positive tests on 574 individuals up until March 31st. The majority of positive cases were admitted to hospital (483/574, 84%, Table S4), consistent with national recommendations to only test those requiring hospital treatment. Cases detected on admission to hospital had a median reported symptom history of 7 days (interquartile range IQR 3-10, available for 385/483, 80%, of cases; Table S4).

New positive cases increased rapidly to a peak between March 31st and April 8th, before falling steadily through to the end of April (Figure 1). The number of daily new combined probable and definite nosocomial cases initially increased in line with community diagnoses, but peaked a week earlier than hospital SARS-CoV-2 admissions on March 23rd to a maximum of 12 new cases per day. Nosocomial cases then rapidly declined to low levels of 0-2 cases a day during April (Figure 1) and none in the following 4 months (data not shown). When the symptom onset date for community cases admitted to hospital is considered, the peak of symptom onset for community and nosocomial cases overlap (Supplemental Figure 1).

**FIGURE 1.**
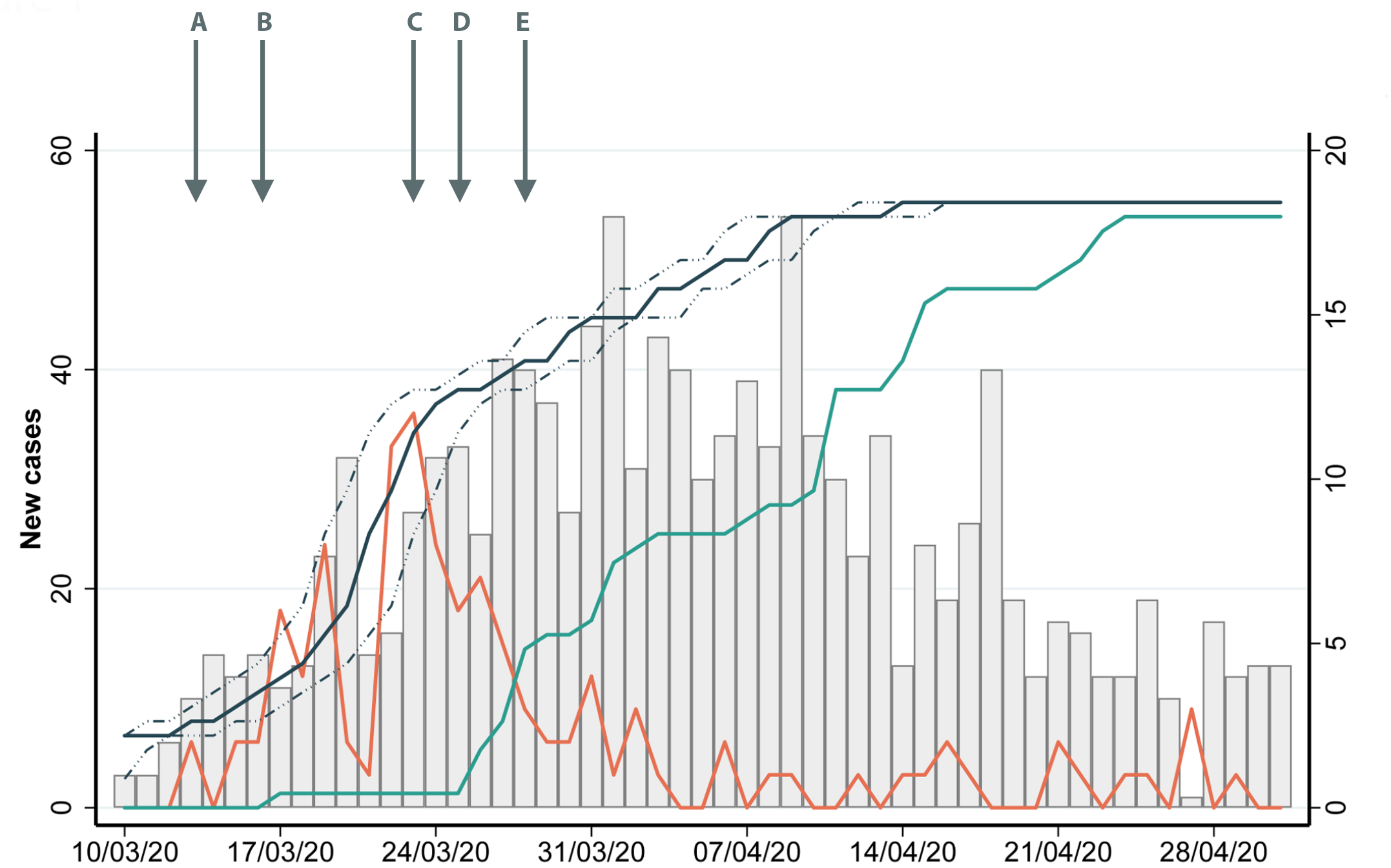
Epidemiological description of cases diagnosed during the first wave. On the left hand y-axis, the grey bar chart displays new cases over time between March 10th and April 31st. Over the same period the right hand y axis shows incidence of nosocomial cases (orange line) and, the proportion (%) of screened HCW with confirmed infection reporting symptom onset (black line) with peak period of infectivity ± 2 days (dashed black line), with IgG seroprevalence of HCW (green). Overlaid is 5 key dates in public policy and infection control (A) March 13th; testing recommended for all inpatients with cough and fever. (B) March 16th; strong government advice for social distancing; (C) March 23rd; implementation of national lockdown (D) March 25th; exclusion of hospital visitors (E) March 28th; mandatory use of surgical masks for all patient interactions under 2 metres.

462 (80%) of the 574 SARS-CoV-2 positive patients were categorised based on NHS England and ECDC definitions (26) as community-onset, with 63 (11%) definite nosocomial, 27 (5%) probable nosocomial and 22 (4%) indeterminate cases (see Methods). Demographics of each group are shown in Table 1. 323 (56%) were male and the median age was 61, although indeterminate, probable nosocomial, and definite nosocomial cases were older (73 years for each group) than community cases (58 years). 225 (49%) of the 455 patients having ethnicity recorded were of black and minority ethnicities (BAME). This proportion was higher amongst community cases (201/371, 54%), compared to indeterminate (6/12, 50%), probable (5/20, 25%) and definite nosocomial cases (13/52, 25%). Comorbidity was lower in community-onset cases 89compared to probable and definite nosocomial cases (median Charlson score 2 (IQR 0-5) versus 5 (IQR 4-6) respectively). The overall crude in-hospital mortality was 20%, ranging from 17% for community-onset cases to 37% for definite nosocomial cases. Of the 574 cases diagnosed until March 31st, 541 were within the study period for genomic analysis between March 13th and 31st. SARS-CoV-2 genome sequence was obtained from 370 of 541 (68%) cases, including 89% (80/90) of all probable and definite nosocomial cases and 72% (168/234) of cases placed into epidemiological clusters (Table S4, S7).

**TABLE 1.** Demographics of the 574 cases diagnosed by the diagnostic lab until March 31st, separated by community-onset, indeterminate, probable nosocomial, and definite nosocomial infections.

|  | Overall |  |  | Community |  | Indeterminate |  | Probable nosocomial |  | Definite nosocomial |  |
| --- | --- | --- | --- | --- | --- | --- | --- | --- | --- | --- | --- |
| Cases (n) | 574 |  |  | 462 | 80% | 22 | 4% | 27 | 5% | 63 | 11% |
| In-hospital mortality | 114 | 20% |  | 78 | 17% | 5 | 23% | 9 | 33% | 23 | 37% |
| Sex |  |  |  |  |  |  |  |  |  |  |  |
| Female | 251 | 44% |  | 203 | 44% | 6 | 27% | 12 | 44% | 30 | 48% |
| Male | 323 | 56% |  | 259 | 56% | 16 | 73% | 15 | 56% | 33 | 52% |
| Median age (IQR) | 61 (48-76) |  |  | 58 (45-73) |  | 73 (61-77) |  | 73 (65-81) |  | 73 (61-81) |  |
| Ethnicity |  |  |  |  |  |  |  |  |  |  |  |
| Asian | 27 | 5% |  | 23 | 5% | 0 | 0% | 1 | 4% | 3 | 5% |
| Black | 157 | 27% |  | 141 | 31% | 4 | 18% | 4 | 15% | 8 | 13% |
| Other | 36 | 6% |  | 33 | 7% | 1 | 5% | 0 | 0% | 2 | 3% |
| White | 230 | 40% |  | 170 | 37% | 6 | 27% | 15 | 56% | 39 | 62% |
| Mixed | 5 | 1% |  | 4 | 1% | 1 | 5% | 0 | 0% | 0 | 0% |
| Unknown | 119 | 21% |  | 91 | 20% | 10 | 45% | 7 | 26% | 11 | 17% |
| Known | 455 | 79% |  | 371 | 80% | 12 | 55% | 20 | 74% | 52 | 83% |
| BAME | 225 | 49% |  | 201 | 54% | 6 | 50% | 5 | 25% | 13 | 25% |
| Pregnant | 13 | 2% |  | 13 | 3% | 0 | 0% | 0 |  | 0 | 0% |
| Smoker |  |  |  |  |  |  |  |  |  |  |  |
| Never | 449 | 78% |  | 373 | 80% | 17 | 77% | 15 | 56% | 44 | 70% |
| Current | 26 | 5% |  | 17 | 4% | 2 | 9% | 3 | 11% | 4 | 6% |
| Ex | 99 | 17% |  | 72 | 16% | 3 | 14% | 9 | 33% | 15 | 24% |
| Alcohol excess | 17 | 3% |  | 12 | 3% | 1 | 5% | 1 | 4% | 3 | 5% |
| Charlson score (IQR) | 3 (1-5) |  |  | 2 (0-5) |  | 4 (3-6) |  | 5 (4-6) |  | 5 (4-6) |  |
| Solid tumor | 71 | 12% |  | 45 | 10% | 6 | 27% | 4 | 15% | 16 | 25% |
| Localised | 53 | 9% |  | 36 | 8% | 3 | 14% | 2 | 7% | 12 | 19% |
| Metastatic | 18 | 3% |  | 9 | 2% | 3 | 14% | 2 | 7% | 4 | 6% |
| Haematological malignancy | 14 | 2% |  | 4 | 1% | 2 | 9% | 2 | 7% | 6 | 10% |
| Lymphoma | 7 | 1% |  | 1 | 0% | 2 | 9% | 1 | 4% | 3 | 5% |
| Leukaemia | 7 | 1% |  | 3 | 1% | 0 | 0% | 1 | 4% | 3 | 5% |
| Renal impairment | 110 | 19% |  | 84 | 18% | 3 | 14% | 9 | 33% | 14 | 22% |
| Mild | 49 | 9% |  | 33 | 7% | 3 | 14% | 4 | 15% | 9 | 14% |
| Moderate | 7 | 1% |  | 5 | 1% | 0 | 0% | 1 | 4% | 1 | 2% |
| Severe | 54 | 9% |  | 46 | 10% | 0 | 0% | 4 | 15% | 4 | 6% |
| Chronic liver disease | 15 | 3% |  | 11 | 2% | 0 | 0% | 0 | 0% | 4 | 6% |
| Dementia | 50 | 9% |  | 29 | 6% | 3 | 14% | 4 | 15% | 14 | 22% |
| Immunosuppression | 35 | 6% |  | 24 | 5% | 0 | 0% | 2 | 7% | 9 | 14% |
| HIV/AIDS | 2 | 0% |  | 2 | 0% | 0 | 0% | 0 | 0% | 0 | 0% |
| Peptic ulcer disease | 9 | 2% |  | 5 | 1% | 0 | 0% | 0 | 0% | 4 | 6% |
| Diabetes mellitus | 168 | 29% |  | 134 | 29% | 5 | 23% | 10 | 37% | 19 | 30% |
| End organ damage | 38 | 7% |  | 27 | 6% | 2 | 9% | 5 | 19% | 4 | 6% |
| Connective tissue disease | 13 | 2% |  | 10 | 2% | 1 | 5% | 0 |  | 2 | 3% |
| Hypertension | 257 | 45% |  | 199 | 43% | 9 | 41% | 17 | 63% | 32 | 51% |
| TIA/CVA | 45 | 8% |  | 28 | 6% | 1 | 5% | 1 | 4% | 15 | 24% |
| Congestive cardiac failure | 28 | 5% |  | 13 | 3% | 0 | 0% | 5 | 19% | 10 | 16% |
| Peripheral vascular disease | 13 | 2% |  | 6 | 1% | 1 | 5% | 3 | 11% | 3 | 5% |
| Myocardial infarction | 19 | 3% |  | 11 | 2% | 1 | 5% | 3 | 11% | 4 | 6% |
| COPD | 46 | 8% |  | 29 | 6% | 2 | 9% | 4 | 15% | 11 | 17% |

### Healthcare worker symptomatology and sero-conversion

The cumulative number of HCWs reporting COVID-19 compatible symptoms and having documented seroconversion from a cohort of 228 HCWs is shown in Figure 1. 43/228 (19%) seroconverted to SARS-CoV-2 IgG, with 44% (19/43) having done so by their first follow-up (April 10th) and 95% (41/43) by May 1st (Table S2). Of note 13/43 (30%) were asymptomatic. Figure 1 presents the predicted period of peak HCW infectectiousness based on a combination of ± 2 days from date of symptom onset (n=30) or seroconversion data where symptoms were not present (n=13). The rapid rise in HCW infection is predicted to have been between March 16th and 25th overlapping with similar rapid rises in community cases reported elsewhere (3), and both symptom reporting by our community-onset cases and incidence of nosocomial cases.

### Linking epidemiology and genomics to define transmission clusters

Strict epidemiological criteria were used to place patients into clusters based on having at least one overlapping time-point in the same ward, outpatient setting, care home or dialysis facility during 14 days prior to their positive sample (Figure 2a). Each cluster had to include a nosocomial case. This identified 44 epidemiological clusters involving 234 cases comprising 123 community admissions, 89 nosocomial acquisitions (62 definite, 27 probable) and 22 indeterminate cases, with a median of 6 patients per cluster (range 2 - 21) (Table S6; Figure 2c for clusters 4-33). Viral genomic haplotype information for each case was then used to resolve the 44 epidemiological clusters to create ‘combined’ final epidemiological and genetic clusters. In combined clusters the majority of all patient haplotypes must be identical, and no patient’s viral haplotype can differ from the main cluster haplotype by more than 1 SNP (Figure 2a). This process resulted in 14 combined epidemiological and genetic clusters (Figure 2b, d). The combined clusters were then mapped onto the original 44 clusters constructed from epidemiological data alone (Figure 2c, Table S8), where each patient in an epidemiological cluster is classified as either belonging to a combined cluster (coloured bar), or having a haplotype that was not in any combined cluster (grey) or without viral sequence (black).

**FIGURE 2.**
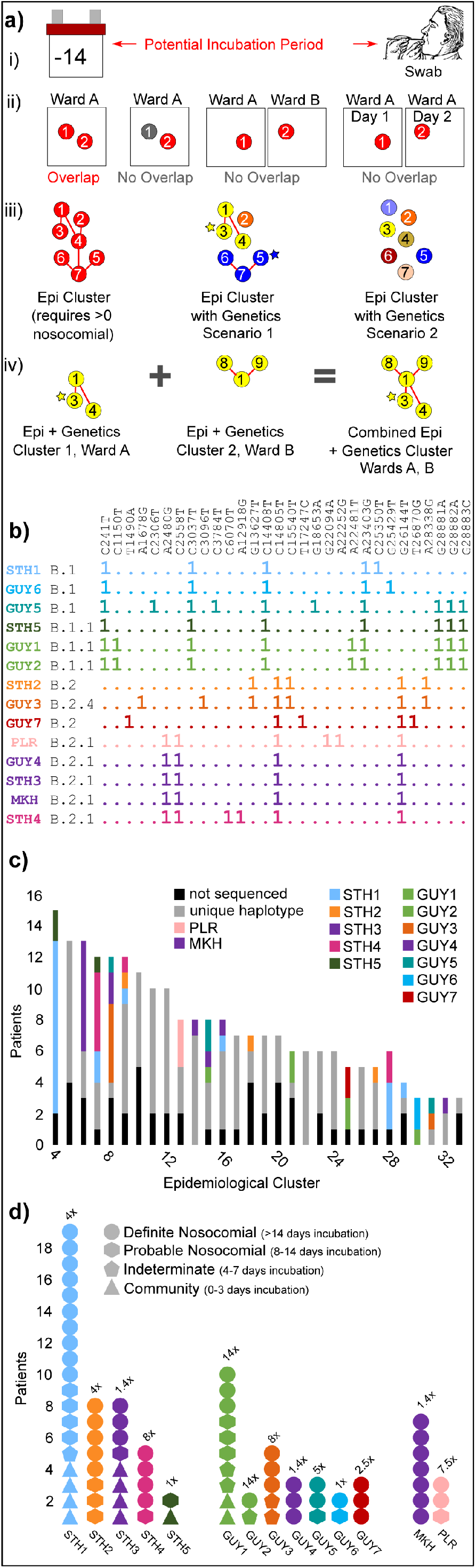
**a)** i) Potential incubation period for a patient is defined as the time from 14 days prior to sample collection up to sample collection. ii) An overlap between two patients (circles with numbers indicating different patients) is defined as two patients who stay on the same ward at the same time, both during their potential incubation period (red circles). The following scenarios are not considered to be overlap: one patient is not in their potential incubation period (grey circle); two potentially incubating patients are on two different wards at the same time; two potentially incubating patients are on the same ward at two non-overlapping times (i.e. different days). iii) An epidemiological (epi) cluster is defined as a set of overlapping patients based on ward stays, where overlap is defined as above, and every patient in the set overlaps at least one other patient in the set, and at least one patient in the set is nosocomial (in hospital >7 days before tested for COVID-19 after having shown symptoms). When viral haplotypes information is taken into account for the patients in a single epi cluster, there are two possible results. In scenario 1, the epi cluster is decomposed into several epi + genetic clusters (indicated by non-red colours) where different colours mean different haplotypes, and a star indicates that a patient has a single additional genetic variant (at most) in addition to the variants comprising the coloured haplotype. In scenario 2, the addition of genetic information decomposes the single epidemiological cluster into individual patients with unique haplotypes, not clustering by genomics. iv) Individual epidemiological plus genomics clusters from different wards are merged if they share at least one patient, and every patient in the resulting merged cluster has at most one SNP difference from the main cluster haplotype. **b)** Haplotype representations of the fourteen clusters that emerge after applying the process depicted in part a) to epidemiological and viral genetic data (see Methods). Clusters are named after the hospital site they occur in (leftmost column). Cluster haplotype lineages are shown in black (second column from left). Cluster haplotypes are depicted (rightmost column) with a “1” in a given position indicating the presence of the SNP relative to the reference genome shown above in vertical text, and a “.” indicating its absence (wild-type sequence). Cluster rows are coloured based loosely on the similarity of the cluster haplotypes to one another. This same colour scheme is used to represent specific clusters in subsequent figures. **c)** Epidemiological clusters 4-33, including cases where n>2 (Table S7) are coloured according to how many of their patients belong to a combined epidemiological plus genomics cluster, with the colour indicative of the viral haplotype (Figure 2b). Patients with viral haplotypes not found in any combined cluster are coloured grey, and those patients for which sequence was unavailable are shown in black. Epidemiological cluster number is shown on the x-axis. Epidemiological cluster 1 -3 are not displayed due to their large size. **d)** Combined epidemiological plus genomic clusters from the acute and elective hospital sites. Clusters are coloured according to viral genomic haplotype (Figure 2b). Clusters are shown broken down into PHE patient nosocomial categories, with different shapes indicating the different categories. Enrichment of the cluster viral haplotype frequency in our study dataset vs. the frequency in the community (Table S8, Methods) is shown on top of each cluster column.

In this manner, 31/44 (70%) epidemiological clusters, including some of the largest, did not have genetic support as they did not have even two cases belonging to any combined cluster (Figure 2c, Table S7). 13/44 (30%) included at least one case from two or more combined clusters, indicating cases from multiple contemporaneous outbreaks harboured within an epidemiological cluster (Figure 2c, Table S7).

### Cluster spatial distribution and enrichment

Of the 14 combined epidemiological and genetic clusters, 5 were on the acute admitting hospital site, 7 were on the elective hospital site and two at intermediate care facilities. These 14 clusters contained 63/80 (79%) of all sequenced probable and definite nosocomial cases (Figure 2d). 3/5 (60%) clusters on the acute site (STH) included a community-onset or indeterminate case, which plausibly served as the originator of SARS-CoV-2 outbreak. This contrasts with the 9 clusters on elective and intermediate sites, which by policy did not knowingly admit COVID-19 patients, where only three (3/9, 33%) included a community-onset or indeterminate case that could have plausibly originated the outbreak (Figure 2d; Supplemental Figure 2 for ward movements). The validity of creating these final 14 clusters was supported by comparing their lineage enrichment compared to community sequences reported to COG-UK consortium CLIMB database during the study period (27). Clusters were enriched by between 1 (for the smallest clusters) and 14-fold (median 4-fold) (Figure 2d, Table S7). In terms of spatial distribution, 7/12 hospital clusters were contained within single wards, 3 clusters were spread on two wards (in 2 cases this involved adjacent wards), and 2 clusters were spread across more than two wards (Figure 4).

**FIGURE 3.**
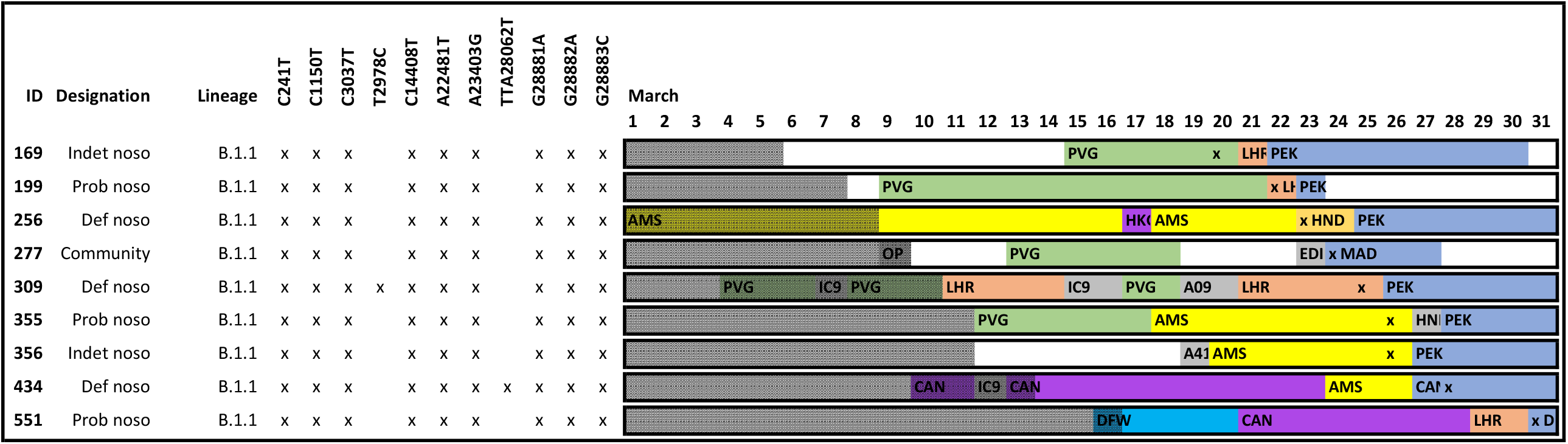
Pictorial representation of ward stays and movements for patients within cluster GUY1. Each row represents a different case. Patient ID, designation, lineage and SNP variants are marked. Ward movements between March 1st to 31st are displayed. Different wards are by given colours. Where there is >1 ward stay on one day, the longest ward stay is represented. The sample collection date is marked with an ‘x’. Time periods outside of the acquisition period are shaded.

**FIGURE 4.**
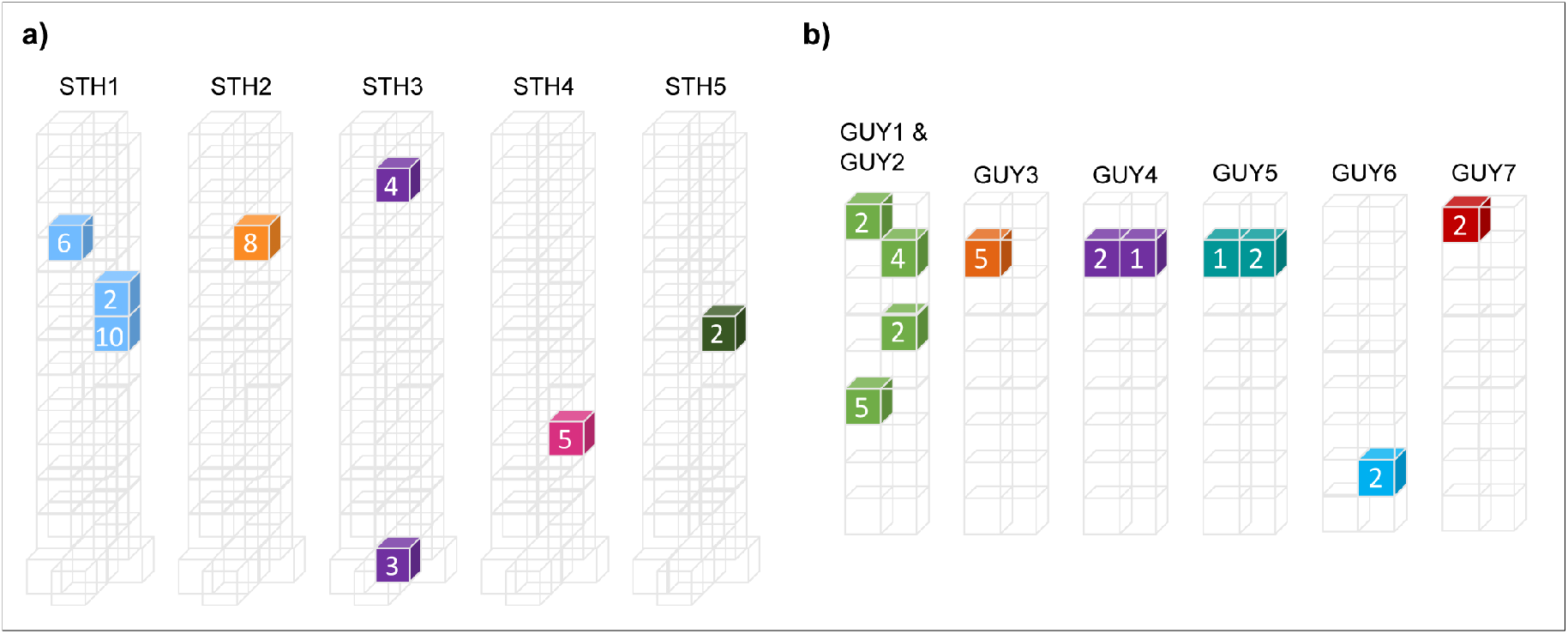
3D spatial representation of St. Thomas’ Hospital **(a)** and Guy’s Hospital **(b)** is shown with wards where transmission of clusters STH1-5 and GUY1-7 occurred (Table S10) are coloured according to viral genetic haplotype (Figure 2b). The numbers in the ward indicate the number of patients from the given cluster inside that ward during their incubation period (as defined in Figure 2a).

### Unsequenced community-onset cases are unlikely to be originators of clusters

We assessed the likelihood that unsequenced community-onset cases may have served as the originator of clusters, where no community-onset or indeterminate cases could be found, by reviewing cases epidemiologically linked with nosocomial clusters. We excluded cases as potential originators if i) they were diagnosed after the first nosocomial case, or ii) available sequence did not meet cluster inclusion criteria (i.e differed from main cluster viral haplotype by >1 SNP), or iii) if they were not community-onset or indeterminate cases (Table S11). Of all the clusters without originators, only 2 clusters could have potentially been originated by an unsequenced community-onset case with epidemiological linkage (case 50 or case 187 for GUY4; case 62 for GUY5).

### Deducing Transmission Networks and Estimating Incubation Period

Patient movements, timing of positive SARS-CoV-2 test, viral haplotype data and SNP acquisition were combined and analysed to infer the place and time period of SARS-CoV-2 acquisition, and predict the direction of transmission from donor to recipient. This analysis could be applied to 5 hospital clusters comprising 24 patients (Figure 5, Supplemental Figure S 3A-D, Table S9 and S10) with patients assigned a lower bound (n=12) and/or upper bound (n=23) for acquisition of SARS-CoV-2, and hence their incubation period (Figure 5a,b,c). The aggregate analyses gave a median lower bound of 6 days and median upper bound of 9 days (Figure 5d, Supplemental Figure 3 A-D, Table S10), consistent with estimates that range from 2-14 days with a median estimate of 5-7 days (28).

**FIGURE 5.**
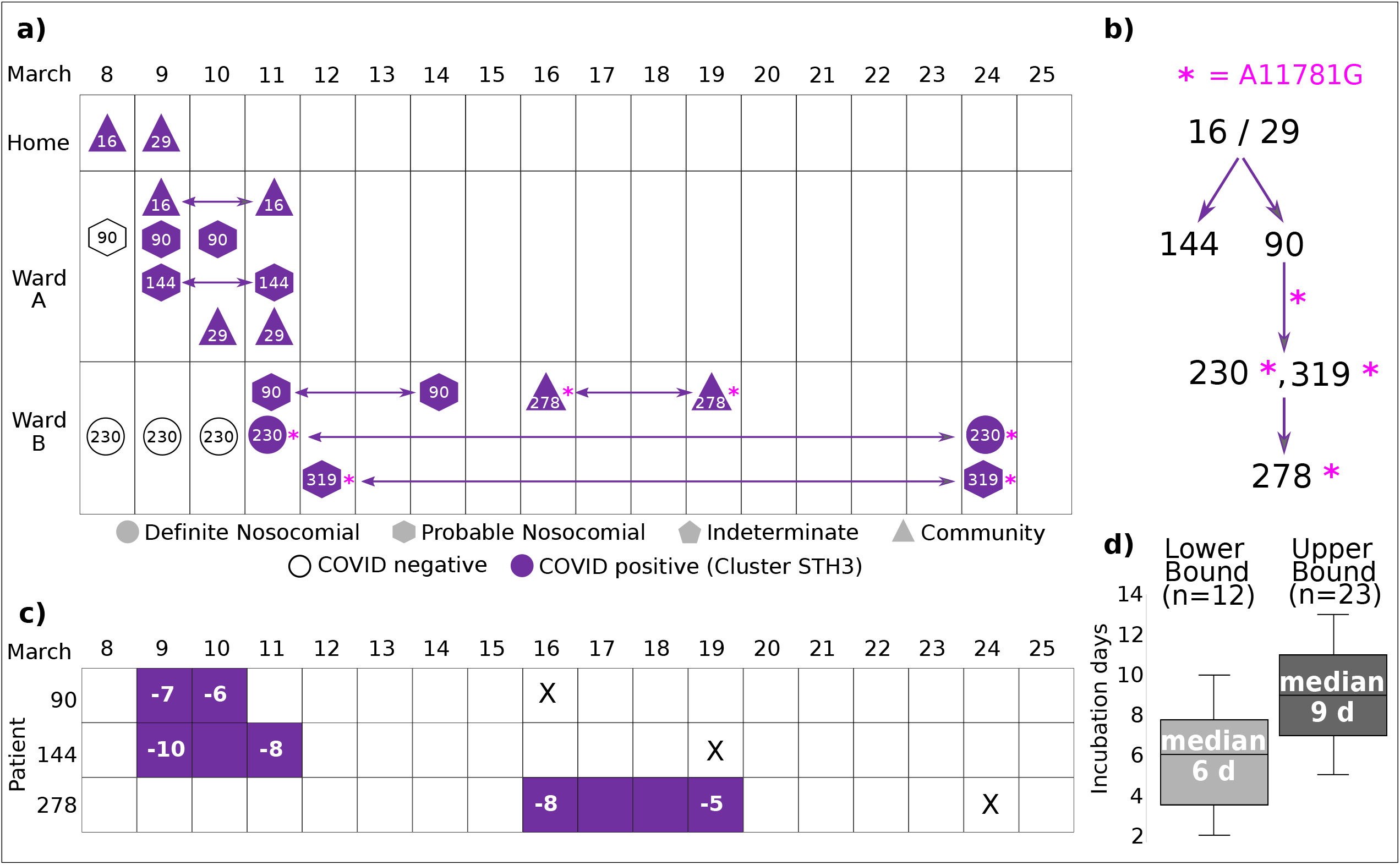
**a)** Patient movement diagram and inferred transmission network for cluster STH3. Patient movements from home to hospital and between wards within St. Thomas’ Hospital are shown from the 8th to the 25th of March. Shapes indicate the nosocomial designation assigned to the patient as in Figure 2 (d). Patients are coloured according to viral haplotype (Figure 2b) from the earliest date at which it can be deduced that they could have caught COVID-19 onwards, based on the viral haplotype and the movement of other patients within the cluster. Prior to the inferred earliest possible infection start date they are shown as white shapes. Double-sided arrows indicate a patient staying on a ward for several days. Patients 16 and 29 originate the cluster, arriving from home sites within 1km of each other on the 8th and 9th, into Ward A (LON), where they transmit the same viral haplotype to patients 90 and 144. Patient 90 is then moved to Ward B (INV) on the 11th, which leads to the infection of patients 230 and 319, and in the process an accumulation of an extra SNP in the viral haplotype (pink asterisk, A11781G). Patient 278 is infected with this new viral haplotype sometime between the 16th and 18th, when they are on ward B. Patient 278 went back and forth between home and the hospital several times, so while strictly designated as “Community”, likely caught this rare hospital-enriched SARS-CoV-2 haplotype in Ward B. **b)** Inferred transmission network. The most parsimonious COVID-19 transmission network for this cluster based on patient ward movements and viral haplotypes is shown. The pink asterisk indicates the mutation of the original haplotype via the acquisition of an additional SNP A11781G. **c)** Inferred lower and upper incubation period bounds for selected patients from cluster STH4. Based on the patient ward movements (part a) and the transmission network (part b), the lower and upper bounds of when certain patients must have contracted SARS-CoV-2 are indicated by coloured squares, with days prior to SARS-CoV-2 test (indicated with an “X”) shown as negative integers. **d)** Aggregate boxplots of lower and upper bounds for incubation period for all patients for which the types of deduction shown in parts c) are possible, for all combined epi + genetic clusters STH1-5, GUY1-7 (Figure S2, Table S11). The number of patients for which the lower bound or upper bound inference was possible is indicated above each boxplot. The median number of days is indicated inside the boxplot for the lower bound and the upper bound for the incubation period.

### Representation of healthcare workers in transmission networks

HCW were required to self isolate immediately upon developing COVID-19 compatible symptoms and were not routinely offered SARS-CoV-2 RNA testing. For this reason there are only 20 SARS-CoV-2 sequences from HCW in the dataset. 17 did not share genomic similarity with other cases, which excludes them from contributing to transmission within sequenced clusters. The viral haplotypes from 3 HCW shared genetic similarity with cluster cases, 1 of which (case 280) cared for a nosocomial case (case 60) within cluster STH1 and shared the same haplotype. Case 280 therefore can be added to cluster STH1, however 280 is unlikely to be the originator of this cluster as they were diagnosed more than 7 days after the first case in this cluster.

## DISCUSSION

This combined epidemiological and genomic analysis spans 19 days of intense community transmission in London during the first wave of the pandemic, which coincided with the main period of nosocomial transmission, comprising 90 definite or probable nosocomial cases across our healthcare institution. This was also the period when stepwise increases in public health restrictions were introduced, including repeated instructions to stay at home and socially distance by at least 2 metres in public places from 16th March, through to national lockdown on March 23rd.

Our transmission analysis during this important period provides evidence informing methodological approaches to nosocomial SARS-CoV-2 outbreak detection. First, both epidemiological and genomic information is essential for accurate linkage of nosocomial cases into putative SARS-CoV-2 clusters: we could not find genomic support for over half of the transmission clusters discovered using epidemiological data alone, and the rest are significantly reduced in size (Fig 3c, Table S7, S8). This latter point is illustrated by epidemiological cluster 8, where genome sequence identified the presence of 3 genomic clusters within a single epidemiologically defined cluster. Second, we show that isolates with a single SNP difference can reasonably be included within clusters given the known SARS-CoV-2 mutation rate and the timeframe of nosocomial transmission clusters. These single SNPs were highly informative as they helped determine the sequence of transmission (Figure 5, Supplemental Figure 3 A-D). Third, it is important to assess the likelihood that linked nosocomial cases even with identical genomes might be due to chance due to the high prevalence of that sequence in the community. We included a method to assess enrichment compared with the surrounding London population by comparing data from the COG-UK CLIMB database, which showed cluster enrichment ranging from 1 to 14 fold. Importantly, the lowest enrichment was seen for clusters of size 2 (Figure 2d). For cases where the enrichment is over 1, this approach gives greater confidence in the definition of clusters and is an important measure that has not thus far been used in other genetic studies. Fourth, we show that for SARS-CoV-2, given its relatively long incubation time, patients should be considered in clusters based on contact during previous admissions. Five patients across 3 clusters (case 520, 362, 236 in STH1, 278 in STH3 and 277 in GUY1, see Supplemental Figure 1) had epidemiological contact during a previous admission to the one during which they were diagnosed. Such contact is not currently considered in published definitions. Finally, we were also able to robustly estimate median incubation period for nosocomial cases as between 6-9 days, which, to our knowledge, is the first time the COVID-19 incubation period has been estimated using both genetics and epidemiological data. This is consistent with published evidence that most people become symptomatic within 7 days of exposure (13). In addition, our study showed a similar proportion of probable and definite nosocomial cases could be placed into transmission clusters. Thus when a probable nosocomial case is found it is likely to be a genuine nosocomial case and should be treated as such.

A number of clusters (GUY1, GUY2, GUY4, GUY5) involved cryptic transmission across multiple wards which is consistent with untested individuals such as staff or asymptomatic patients, but not patient visitors, as missing links in transmission clusters. For instance, clusters GUY1 and GUY2 involved 4 wards, with an identical viral haplotype circulating with high enrichment compared to the community. Whilst these two clusters meet our cluster definition as defined in Methods, they are not merged due to there being no patient overlap in ward stay between the clusters. Given these were highly enriched clusters, in the same building just a few floors apart, and with no community-onset case as the originator of GUY2, the most parsimonious explanation is cryptic transmission between the two. Similarly in GUY1 there is no patient who clearly could have infected case #551 (see Supplemental Figure 1). Lastly, GUY4 and GUY5 involve highly enriched haplotypes across neighbouring wards. These situations suggest cryptic transmission, and ‘missing links’ which may be HCW or asymptomatic, untested patients.

With the above methodological approaches defined, we focussed on the initial period of nosocomial SARS-CoV-2 transmission to try and identify factors explaining its rapid rise and fall. There was initially a complete temporal overlap between the increase in community cases, healthcare worker symptoms, and incidence of nosocomial cases. We propose that the most likely explanation is that HCWs were an essential contributor to the intensity of nosocomial transmission during this initial period, potentially by both initiating and sustaining transmission. This is supported by a number of findings. First, reported symptoms in HCW with confirmed infection were before or at the time of lockdown on 23rd March, with their peak infectivity predicted to coincide with peak nosocomial transmission (Figure 1). Second, cryptic transmission between wards was clearly demonstrated, suggesting involvement of untested individuals moving between wards, with HCWs being the most likely vector. Third, we were unable to identify community or indeterminate cases who may have served as originators for more than half of clusters, suggesting involvement of untested individuals such as HCW. Indeed many clusters also occurred on the elective sites, where cases were not knowingly admitted. Finally, we did link 1 of only 20 tested HCWs to a transmission cluster, which although not the originator, is consistent with the potential for HCWs to sustain outbreaks. Whilst HCW seem the most likely source of transmission it is however possible that unrecognised patient-to-patient transmission occurred, for instance by asymptomatic patients or in areas where we do not have epidemiological data, such as radiology departments

Subsequently, from around March 16th, progressive CSD rapidly reduced community SARS-CoV-2 transmission, which would also decrease the incidence of HCWs infected in the community, lowering their potential to contribute to nosocomial transmission. Consistent with this conclusion, nosocomial infection fell rapidly from March 23rd to the end of the study in line with reported community infections, and before any potential impact from surgical mask use or banning of visitors to hospital.

It is harder to interpret events after 31st March. It is notable that SARS-CoV-2 patients continued to be admitted, peaking on 8th April, whilst nosocomial cases fell and remained low. One conclusion is that admission of community-onset cases is in itself unlikely to cause large clusters. Partly this may be due to rapid diagnosis of clinically suspected cases and pre-emptive isolation, in an environment where HCWs are wearing appropriate PPE. Additionally, It may be that by the time community-onset cases of SARS-CoV-2 are admitted to hospital (in our study a median day 7 days post onset of symptoms) they are less infectious because viral load is falling. Indeed, this is consistent with previous studies showing a median duration of viral shedding of 4 days post onset of symptoms (29).

In summary, this study supports routine use of genome sequencing for SARS-CoV-2 outbreak investigation and provides a framework for data interpretation that calls into question continued reliance on using epidemiology alone. A multi-site clinical trial, COG-UK HOCI, is currently investigating whether provision of real-time sequencing for outbreak analysis can help infection control teams interrupt transmission (30). We provide data supporting a conclusion that HCWs infected in the community are a major contributor to nosocomial transmission during the initial stage of the first wave of the pandemic, which was halted by implementation of CSD policies. With community transmission returning, including to HCWs, there will be an opportunity to assess whether nosocomial transmission returns to the same intensity with policies such as universal mask use in place, or whether additional interventions such as regular staff testing or rapid SARS-CoV-2 genome sequencing is required to limit nosocomial transmission. This is particularly important given the challenges re-introducing CSD policies and the need to continue with non-COVID-19 clinical activity.

## METHODS

### Setting

Guy’s and St Thomas’ Hospital NHS Foundation Trust includes: 1) an acute admitting adult hospital site with an Emergency Department (St Thomas’) also hosting the network lead centre for nationally-commissioned respiratory high consequence infectious diseases (HCID), which admitted some of the first UK SARS-CoV-2 cases to specialised intensive care and general wards. It was also the only site knowingly admitting COVDI-19 patients. 2) an elective ambulatory site including elective orthopaedic and thoracic surgery, a cancer centre, dialysis and renal transplant unit (Guy’s); 3) two long stay community care units and 4) multiple community dialysis units. All sites are part of the same management structure and were part of a single operational pandemic response structure with daily meetings throughout and received the same laboratory diagnostic and infection control services. Infection control policies were reviewed daily with reference to frequent updates from Public Health England and NHS England, to implement stepwise measured derogation of Personal Protective Equipment (PPE) use while caring for known and suspected COVID-19 patients moving from HCID designation to caring for hundreds of COVID-19 patients. Hospital visitors and relatives were excluded from the hospital from March 25th, and universal HCW surgical mask use policy was introduced for all patient interactions across all hospital and community facilities on March 28th.

### Sample retrieval and clinical data collection

490 surplus RNA extracts from SARS-CoV-2 combined nose and throat swabs collected between 13th and 31st March 2020 inclusive, predominantly from patients within the catchment South East London boroughs, and stored in the routine diagnostic laboratory after completion of all clinically-requested tests, were retrieved by the primary care team. Swabs had been tested using multiplexed tandem PCR (AusDiagnostics two-step multiplexed-tandem (MT-PCR) Coronavirus Typing Eight-well Panel; cat. no. 2061901) targeting ORF1a of SARS-CoV-2. 448 extracts had been reported as SARS-CoV2 detected and 42 not-detected. The cycle threshold (Ct) value was taken as 15 plus the diagnostic MT-PCR take off value. Isolates with a Ct of 16 – 37 were regarded as eligible for WGS. Extracts were anonymised and linked with a limited set of clinical data (Table 1) and the admission and ward stay dates by the primary care team, before submission to the research team. Favourable opinion to conduct this work was granted by the North West Preston Research Ethics Committee (Reference 18/NW/0584).

### Anonymisation

All patients and samples were anonymised using a unique patient and sample identification number. Ward stays were anonymised using the following schema: locations beginning with ‘A’ represent ambulatory or outpatient locations numbered A01 to A60; if prefixed with ‘ED’ represent emergency departments; if starting with ‘IC’ represent an intensive care unit; each patients usual residence was named ‘Home’ followed by their anonymised patient ID, and inpatient wards were anonymised using three letter international airport codes.

### COVID-19 Whole Genome Sequencing

SARS-CoV-2 positive RNA isolates were processed as described in pcr-tiling-ncov-PTC_9096_v109_revF_06Feb2020-gridion from Oxford Nanopore Technologies (ONT) with V3 PCR primers from the ARTIC protocol (31). 24 samples were prepared in each library. 3µl of synthesised cDNA was used in each PCR reaction instead of the ONT suggested 2.5µl and 35 PCR cycles were conducted. The 20°C incubation was extended to 10 minutes in the end-prep step. All the AMPure XP bead clean-up steps used a 1:1 ratio of library to AMPureXP beads. The post-barcode ligation bead clean-up was washed with 2x 450µl SFB washes rather than the ONT suggested 700µl. 17.5ng of the final library was loaded onto the flow cell for sequencing. The flow cell sequenced for 3 days or until there were very few active pores remaining.

### Generation of SARS-CoV-2 consensus sequence and variant and lineage calling from raw Nanopore reads

From raw Nanopore reads, COVID-19 viral consensus sequences and variants were generated using the Artic bioinformatic pipeline v 1.0 as described in Artic Network(32). Lineages were assigned using the Pangolin software (33) version 1.1.14 with lineages version 2020-05-19.

### Haplotype construction for patients

For each patient, a viral haplotype was constructed by juxtaposing all variants from the .pass.vcf file output by the Artic bioinformatics pipeline above. The exception was for genomic positions that were known to be homoplasic according to problematic sites (https://raw.githubusercontent.com/W-L/ProblematicSites_SARS-CoV-2/master/problematic_sites_sarsCov2.vcf). These sites at positions 635, 11074, 11083, 16887, 21575, were all masked and not included in any haplotypes. For patient 363, three variants (C3096T, G13627T, C15540T) were also taken from the .fail.vcf file. This patient was in a transmission cluster with strong epidemiological evidence linking them to 4 patients who all had variants C3096T, G13627T, and C15540T. All three of these variants were found in all of the other patients in this transmission cluster. This situation with variants in the .fail.vcf file rather than the .pass.vcf file only arose for patient 363 in our dataset.

### Definitions and deduction of transmission Clusters

Transmission clusters were deduced in a series of stages using combined epidemiological and genetic information, in three phases as follows. First, each patient sample was placed in nosocomial categories based on date of testing according to NHS England and ECDC definitions (26): Community (<3 days from admission), indeterminate (3-7 days from admission), probable Nosocomial (8-14 days from admission) and definite nosocomial (>14 days from admission). Epidemiological clusters were constructed using the following rules (Fig 2a and table S7): At least two cases in the epidemiologically defined cluster; at least one probable or definite nosocomial case; all cases in the seed cluster must overlap with at least one other patient in the same seed cluster. Overlap was defined as both patients being on the same ward at the same time for at least one day during the incubation period set as 14 days prior to the positive SARS-CoV-2 positive swab (13).

In the second phase of cluster construction, genetic information was added by considering the viral haplotype for each patient in the epidemiological cluster. Epidemiological clusters were split into combined epidemiological + genetic clusters where every patient has the same viral haplotype. These combined epidemiological plus genetic clusters are termed “seed” clusters.

In the third phase, additional patients were added to seed clusters based on the following criteria: the patient added must overlap with at least one patient in the seed cluster (where overlap is defined as above); the added patient has a viral haplotype that differs only one SNP from the seed cluster viral haplotype. Patients who met criterion 1 (overlap) but whose viral haplotypes differed by 2 SNPs from the seed cluster viral haplotype were added to the cluster if at least one of the “missing” SNP positions was called as an N due to insufficient sequence coverage in consensus sequence generation. For two clusters, (STH4, STH5) phase 3 involved merging two different seed clusters whose viral haplotypes differed from each other by one SNP.

There are two clusters that are exceptions to the above rules: GUY3 and GUY4. These two clusters were added upon manual inspection of the combined epidemiological and genetic data. In both cases, two patients whose viral haplotype differed by a single SNP overlapped on the same ward during their incubation period, and a third patient, whose viral haplotype was identical to that of one of the two previously mentioned patients was present on an adjacent ward at the same time, also during their incubation period. These two clusters violate the criterion given above that seed clusters should be on a single ward, but are nonetheless considered as transmission clusters because the two wards (Samaritan and Hedley Atkins) in both cases are spatially adjacent and with frequent sharing of staff across wards, especially at night..These clusters also showed high enrichment compared to community haplotypes (see below.)

### Calculation of Hospital compared with Community Enrichment for cluster viral haplotypes

Enrichment of viral haplotype frequencies amongst nosocomial and community cases were calculated as follows (Table S8 and Fig 2). For a given haplotype, the hospital frequency (H) was calculated by dividing the number of patients in the dataset by the total number of sequenced patients in our dataset (370). COG-UK consortium data in COV_GLUE was extracted on 29th June 2020 to assess community haplotype frequencies (27). To assist with SQL queries, a view from several tables was created with the following SQL query: “create table full as select m.id as sequence_id,m.collection_date as collection_date,adm0,adm1,adm2,num_ns,longest_n_run,num_unique_snps,pang_lineage,cov_glue_lineag e,cov_nt_mutation_id as mut_id from cov_coguk_metadata m join cov_sequence s join cov_cov_nt_mutation_sequence ms on m.id = sequence_id and ms.sequence_sequence_id = sequence_id ;”

Two numbers were used to calculate the community frequency,: the entire community population of relevance, and the number of patients with the given haplotype in the community population. The community population (Cd) was defined as all the sequenced patients within the study period (13th to 31st March inclusive) with a location of ‘GREATER LONDON’, ‘LONDON’, or ‘CITY OF LONDON’, whichever had greater than 90% sequence coverage (952 people). This number was obtained using the following SQL query: “select count(distinct sequence_id) from full where (adm2 like ‘%LONDON%’ and adm2 != ‘LONDONDERRY’) and (collection_date >= ‘2020-03-13’ and collection_date <= ‘2020-03-31’) and num_ns/29401 < 10;” Where 29401 is the length of the reference viral genome coding region, so given the percentage of Ns in the sequenced sample. Requiring a cutoff of less than 10% Ns is equivalent to greater than 90% sequencing coverage, which is the cutoff required for our samples in this study.

The number of patients with the given haplotype in the community population was calculated by determining the number of sequences in the community population that had all of the SNP variants in the given haplotype, and no more. For this purpose, any homoplasic SNPs that had been removed from consideration when constructing patient viral haplotypes within our study population (see section “*Haplotype construction for patients*” above), namely SNPs at positions 635, 11074, 11083, 16887, 21575, were added back to calculate the haplotype numbers in the community, as these SNPs had not been removed in the samples submitted to COG-UK. The following is an example query to obtain the number of patients in the community with the haplotype for our cluster STH1 (C241T,C3037T,C14408T,A23403G,C25350T):

“select distinct sequence_id from full where 100*(num_Ns/29903) < 10 and sequence_id in (select sequence_id from full where collection_date >= ‘2020-03-13’ and collection_date <= ‘2020-03-31’ and mut_id = ‘C3037T’) and sequence_id in (select sequence_id from full where collection_date >= ‘2020-03-13’ and collection_date <= ‘2020-03-31’ and mut_id = ‘C14408T’) and sequence_id in (select sequence_id from full where collection_date >= ‘2020-03-13’ and collection_date <= ‘2020-03-31’ and mut_id = ‘A23403G’) and sequence_id in (select sequence_id from full where collection_date >= ‘2020-03-13’ and collection_date <= ‘2020-03-31’ and mut_id = ‘C25350T’) and (adm2 like ‘%LONDON%’ and adm2 != ‘LONDONDERRY’) group by sequence_id having count(distinct mut_id) = 4”

Results were obtained for 11 community sequences where community frequency was defined as Cn / Cd. A correction factor of 1 was then added to Cn as a pseudocount to avoid dividing by zero for cases where Cn = 0. Thus the formula for community frequency is (Cn + 1) / Cd and the formula to calculate nosocomial enrichment for a given haplotype was H / [(Cn + 1) / Cd].

### Healthcare worker (HCW) serology and symptom reporting

Blood sample collection to determine COVID-19 seroconversion was offered to consenting healthcare workers at Guy’s & St Thomas’ Hospital after expedited approval from the Trust’s R&D office, occupational health and medical director ((34)). Sequential serum samples were collected approximately every 1-2 weeks from 228 HCWs between 13^th^ March and 10^th^ June 2020. Samples were tested using a published ELISA assay with sera diluted 1:50 considered positive if they gave an OD for IgG against both N and S that was 4-fold above the negative control sera. Self-reported COVID-19 related symptoms were recorded by participants and days post onset of symptoms in seropositive individuals was determined using this information. For asymptomatic seropositive individuals, days POS was defined as the first timepoint SARS-CoV-2 Abs were detected. Six HCWs had a confirmed SARS-CoV-2 PCR+ infection. Serology results were communicated individually to HCWs and collectively to the hospital as part of preparation for universal HCW serology testing that commenced in May 2020.

## Data Availability

Sequence data will be available at ENA
Study accession number is: PRJEB41373
Study name is: ena-STUDY-KING'S COLLEGE LONDON-17-11-2020-01:31:23:025-468

## SUPPLEMENTAL TABLE LEGENDS

**SUPPLEMENTAL FIGURE 1** Epidemiological description of cases diagnosed during the first wave. On the left hand y-axis, the grey bar chart displays cumulative symptom onset for admitted cases where symptom onset is known, between March 10th and April 31st. Over the same period the right hand y axis shows incidence of nosocomial cases (orange line) and, the proportion of screened HCW with confirmed infection reporting symptom onset (olive line), and IgG seroprevalence of HCW (green).

**SUPPLEMENTAL FIGURE 2** Ward movement diagrams for the combined clusters formed through epidemiological and genetic analysis. Each row represents one patient’s movements. Lineage, nosocomial designation and haplotype are shown. Ward stays are shown coloured by different ward location, with time on the horizontal axis. Where a patient is on two wards in one day, the ward with the longest stay is represented. An ‘x’ marks date of specimen collection for the first positive PCR test for SARS-CoV-2 for each patient.

**SUPPLEMENTAL FIGURES 3A - 3D** Patient movement and transmission network analogous to figure 5 a-c for clusters STH1 (Figure S 3A), STH4 (Figure S 3B), GUY1 (Figure S 3C), and GUY3 (Figure S 3D). Patient shapes are as specified for Figure 5. Colours are for cluster-specific viral haplotypes, as specified in Figure 2b. Ward names are independent for each cluster (so “Ward A” on Figure S 3A is not necessarily the same as “Ward A” on Figure S 3B).

**SUPPLEMENTAL FIGURE 3A** Patient 127 is the likely origin of the part of cluster STH1 that is on Ward A (KUL ward from Table S5, Table S8). Patient 127 arrives from home to the ward where the nosocomial patients are infected (Ward A) within a two day period. Nosocomial patients 245, 206, 354, 293, and 480 are infected in Ward A after the arrival of patient 127 onto Ward A having already been on this ward before patient 127 arrives (245,480, 293, 206) or arriving after patient 127 (354). Patients 245, 480, 293, and 206 are then transferred to Ward C (SFO), which sets a lower bound on when they must have been infected (as they were infected in Ward A). Subsequently nosocomial patient 469 is infected on Ward C. Independently, patient 127 is moved to Ward B (DEL), where patient 268 gets infected.

**SUPPLEMENTAL FIGURE 3B** Two transmission scenarios (i and ii) are considered for cluster STH4. Either patient 212 arrives on Ward A already a carrier, which leads to the infection of nosocomial patients 337, 396, 351, and 374 (scenario “i”), or the viral haplotype was already circulating on Ward A (DEN) prior to the arrival of patient 212 (scnarion “ii”). In scenario (i), patient 212 transmits the virus to one of patients 337, 396, or 351, and in this transmission, SNP A12918 (purple asterisk) is acquired. Subsequently patient 212 transmits the strain without A12918 to patient 374. In scenario (ii), when patient 212 arrives on Ward A, there are already two strains that differ by a single SNP (A12918) circulating on that ward. “X” and “Y” indicate two separate unknown “missing links” -- either asymptomatic patients or staff. For both scenarios i and ii, the inferred upper bounds for acquisition are identical for patients 374, 396, 351, shown in part c.

**SUPPLEMENTAL FIGURE 3C** Patient 277 originates cluster GUY1, arriving from home onto Ward A (PVG), where nosocomial patients 355 and 199 are infected. Nosocomial patient 309 is infected on Ward A, picking up the additional variant TTA28062T. Patient 355 is then transferred to Ward B (AMS), which leads to the infection of nosocomial patient 256 and patient 356. In an independent transmission network, nosocomial patient 434 is infected on Ward C (CAN) after the arrival of patient 551, and in the process acquires SNP T2978C.

**SUPPLEMENTAL FIGURE 3D** Patient 71 originates cluster GUY3, arriving from home onto Ward A (IC9), then moving to Ward B (HKG) within three days, where nosocomial patients 229, 295, and 363 are infected. All three of these patients acquire SNP A1678G in the process, so it is most parsimonious that patient 71 transmitted to one of these patients, who then transmitted it to at least one of the other two. Several days later, patient 301 is admitted to Ward B, where they are infected, and acquire SNP A24033G in the process.

**SUPPLEMENTAL TABLE 1** Number of tests performed, number of positive tests, number of new cases, combined number of probable and definite nosocomial cases, and number of community viral sequences submitted to CLIMB, all by date from March 9th to March 31st.

**SUPPLEMENTAL TABLE 2** Cumulative incidence of HCW IgM and IgG seroconversion by week, beginning on Mar 13th until Jun 12th. Total number sampled (n=228)

**SUPPLEMENTAL TABLE 3** Genomic sequence of viral isolates from cases. Sequence only included if sequenced successfully with >90% coverage at 8x depth. GISAID accession ID, lineage, haplotype, coverage and N positions for each genomic sequence displayed.

**SUPPLEMENTAL TABLE 4** For each patient, symptom onset (column B), hospital admission or outpatient encounter date (column C) during which first positive SARS-CoV-2 samples is taken. Sample collection date and time (column D) is given, with time between symptom onset and admission (column E) or admission/encounter and sample collection date (column F). The admission/encounter is categorised (column G) as either ‘outpatient’, ‘A&E’ where samples are taken in the emergency room when the patient is not admitted, and ‘Inpatient’ where the patient is admitted during that encounter. Column H categorises patients as either as per the NHS England and ECDC definitions of nosocomial infection (26). Column I shows whether viral sequence for the isolate was successfully obtained..

**SUPPLEMENTAL TABLE 5:** Patient ward movements. Every encounter or ward stays in the 14 days prior to the first positive test is given, with start and end date. Each row represents a different encounter or ward stay. Ward stays are arranged by ascending patient number and in chronological order.

**SUPPLEMENTAL TABLE 6:** Epidemiological clusters with ward of overlaps and patients within the cluster. Clusters must contain at least one probable or nosocomial infection (see Methods). Sorted from largest cluster to smallest.

**SUPPLEMENTAL TABLE 7:** Epidemiological clusters separated into those that isolates that were not sequenced successfully, genetic haplotypes without genetic similarity to other sequenced individuals, and by haplotypes that form clusters with other cases (GUY1 to GUY7, STH1 to STH5, MKH, PLR)

**SUPPLEMENTAL TABLE 8:** Final transmission clusters with associated hospital site, ward, ward type, lineage and case numbers. Overall number of each noscomail designation for the clusters is given in column G. Seed haplotype is given, with SNP variants from the seed haplotype also reported. The final three columns show the calculation of the hospital enrichment factor; the number of sequences with the seed haplotype from the sequenced isolates from hospital, the number of community samples with this haplotype submitted to CLIMB, and the enrichment factor which is the ratio of these two numbers.

**SUPPLEMENTAL TABLE 9:** Data used for Fig 3 -- number of patients on each ward for each cluster, for each ward with a provable transmission.

**SUPPLEMENTAL TABLE 10:** Lower and Upper bounds for the incubation period (days before onset of symptoms) for specific patients based on inferred transmission networks from Fig 5, Fig S2.

**SUPPLEMENTAL TABLE 11:** Epidemiological clusters that contribute to combined epi + genetic cluster without community-onset or indeterminate cases are assessed to see whether they could contain non- sequenced cases that may serve as originators to the cluster. Cases are deemed not to represent a potential non-sequenced originator if they are a probable or definite nosocomial case, diagnosed after the start of the cluster, or if they do not share significant genome similarity with other cases in the cluster. Epidemiological cluster information is used from Supplemental Table 7

## APPENDIX

<u>Funding acquisition, leadership, supervision, metadata curation, project administration, samples, logistics,</u>

<u>Sequencing, analysis, and Software and analysis tools:</u>

Dr Thomas R Connor PhD33, 34, and Professor Nicholas J Loman PhD15.

<u>Leadership, supervision, sequencing, analysis, funding acquisition, metadata curation, project administration, samples, logistics, and visualisation:</u>

Dr Samuel C Robson Ph.D 68.

<u>Leadership, supervision, project administration, visualisation, samples, logistics, metadata curation and software and analysis tools:</u>

Dr Tanya Golubchik PhD 27.

<u>Leadership, supervision, metadata curation, project administration, samples, logistics sequencing and analysis:</u>

Dr M. Estee Torok FRCP 8, 10.

<u>Project administration, metadata curation, samples, logistics, sequencing, analysis, and software and analysis tools:</u>

Dr William L Hamilton PhD 8, 10.

<u>Leadership, supervision, samples logistics, project administration, funding acquisition sequencing and analysis:</u>

Dr David Bonsall PhD 27.

<u>Leadership and supervision, sequencing, analysis, funding acquisition, visualisation and software and analysis tools:</u>

Dr Ali R Awan PhD 74.

<u>Leadership and supervision, funding acquisition, sequencing, analysis, metadata curation, samples and logistics:</u>

Dr Sally Corden PhD 33.

<u>Leadership supervision, sequencing analysis, samples, logistics, and metadata curation:</u>

Professor Ian Goodfellow PhD 11.

<u>Leadership, supervision, sequencing, analysis, samples, logistics, and Project administration:</u>

Professor Darren L Smith PhD 60, 61.

<u>Project administration, metadata curation, samples, logistics, sequencing and analysis:</u>

Dr Martin D Curran PhD 14, and Dr Surendra Parmar PhD 14.

<u>Samples, logistics, metadata curation, project administration sequencing and analysis:</u>

Dr James G Shepherd MBChB MRCP 21.

<u>Sequencing, analysis, project administration, metadata curation and software and analysis tools:</u>

Dr Matthew D Parker PhD 38 and Dr Dinesh Aggarwal MRCP 1, 2, 3.

<u>Leadership, supervision, funding acquisition, samples, logistics, and metadata curation:</u>

Dr Catherine Moore33.

<u>Leadership, supervision, metadata curation, samples, logistics, sequencing and analysis:</u>

Dr Derek J Fairley PhD 6, 88, Professor Matthew W Loose PhD 54, and Joanne Watkins MSc 33.

<u>Metadata curation, sequencing, analysis, leadership, supervision and software and analysis tools:</u>

Dr Matthew Bull PhD33, and Dr Sam Nicholls PhD 15.

<u>Leadership, supervision, visualisation, sequencing, analysis and software and analysis tools:</u>

Professor David M Aanensen PhD 1, 30.

<u>Sequencing, analysis, samples, logistics, metadata curation, and visualisation:</u>

Dr Sharon Glaysher 70.

<u>Metadata curation, sequencing, analysis, visualisation, software and analysis tools:</u>

Dr Matthew Bashton PhD 60, and Dr Nicole Pacchiarini PhD 33.

<u>Sequencing, analysis, visualisation, metadata curation, and software and analysis tools:</u>

Dr Anthony P Underwood PhD 1, 30.

<u>Funding acquisition, leadership, supervision and project administration:</u>

Dr Thushan I de Silva PhD 38, and Dr Dennis Wang PhD 38.

<u>Project administration, samples, logistics, leadership and supervision:</u>

Dr Monique Andersson PhD 28, Professor Anoop J Chauhan 70, Dr Mariateresa de Cesare PhD26, Dr Catherine Ludden 1,3, and Dr Tabitha W Mahungu FRCPath 91.

Sequencing, analysis, project administration and metadata curation:

Dr Rebecca Dewar PhD 20, and Martin P McHugh MSc20.

Samples, logistics, metadata curation and project administration:

Dr Natasha G Jesudason MBChB MRCP FRCPath 21, Dr Kathy K Li MBBCh FRCPath 21, Dr Rajiv N Shah BMBS MRCP MSc 21, and Dr Yusri Taha MD, PhD 66.

Leadership, supervision, funding acquisition and metadata curation:

Dr Kate E Templeton PhD 20.

Leadership, supervision, funding acquisition, sequencing and analysis:

Dr Simon Cottrell PhD 33, Dr Justin O’Grady PhD 51, Professor Andrew Rambaut DPhil 19, and Professor Colin P Smith PhD93.

Leadership, supervision, metadata curation, sequencing and analysis:

Professor Matthew T.G. Holden PhD 87, and Professor Emma C Thomson PhD/FRCP 21.

<u>Leadership, supervision, samples, logistics and metadata curation:</u>

Dr Samuel Moses MD 81, 82.

<u>Sequencing, analysis, leadership, supervision, samples and logistics:</u>

Dr Meera Chand 7, Dr Chrystala Constantinidou PhD 71, Professor Alistair C Darby PhD 46, Professor Julian A Hiscox PhD 46, Professor Steve Paterson PhD 46, and Dr Meera Unnikrishnan PhD 71.

<u>Sequencing, analysis, leadership and supervision and software and analysis tools:</u>

Dr Andrew J Page PhD 51, and Dr Erik M Volz PhD96.

<u>Samples, logistics, sequencing, analysis and metadata curation:</u>

Dr Charlotte J Houldcroft PhD 8, Dr Aminu S Jahun PhD 11, Dr James P McKenna PhD 88, Dr Luke W Meredith PhD 11, Dr Andrew Nelson PhD 61, Sarojini Pandey MSc 72, and Dr Gregory R Young PhD 60.

<u>Sequencing, analysis, metadata curation, and software and analysis tools:</u>

Dr Anna Price PhD34, Dr Sara Rey PhD 33, Dr Sunando Roy PhD 41, Dr Ben Temperton Ph.D 49, and Matthew Wyles 38.

<u>Sequencing, analysis, metadata curation and visualisation:</u>

Stefan Rooke MSc19, and Dr Sharif Shaaban PhD87.

<u>Visualisation, sequencing, analysis and software and analysis tools:</u>

Dr Helen Adams PhD 35, Dr Yann Bourgeois Ph.D 69, Dr Katie F Loveson Ph.D 68, »ine O’Toole MSc19, and Richard Stark MSc 71.

<u>Project administration, leadership and supervision:</u>

Dr Ewan M Harrison PhD 1, 3, David Heyburn 33, and Professor Sharon J Peacock 2, 3

<u>Project administration and funding acquisition:</u>

Dr David Buck PhD26, and Michaela John BSc Hons 36

<u>Sequencing, analysis and project administration:</u>

Dorota Jamrozy 1, and Dr Joshua Quick PhD 15

<u>Samples, logistics, and project administration:</u>

Dr Rahul Batra MD78, Katherine L Bellis BSc (Hons) 1, 3, Beth Blane BSc 3, Sophia T Girgis MSc 3, Dr Angie Green PhD 26, Anita Justice MSc 28, Dr Mark Kristiansen PhD 41, and Dr Rachel J Williams PhD 41.

<u>Project administration, software and analysis tools:</u>

Radoslaw Poplawski BSc 15.

<u>Project administration and visualisation:</u>

Dr Garry P Scarlett Ph.D69.

<u>Leadership, supervision, and funding acquisition:</u>

Professor John A Todd PhD 26, Dr Christophe Fraser PhD 27, Professor Judith Breuer MD 40,41, Professor Sergi Castellano PhD 41, Dr Stephen L Michell PhD 49, Professor Dimitris Gramatopoulos PhD, FRCPath73, and Dr Jonathan Edgeworth PhD, FRCPath 78.

<u>Leadership, supervision and metadata curation:</u>

Dr Gemma L Kay PhD 51.

<u>Leadership, supervision, sequencing and analysis:</u>

Dr Ana da Silva Filipe PhD 21, Dr Aaron R Jeffries PhD 49, Dr Sascha Ott PhD 71, Professor Oliver Pybus 24, Professor David L Robertson PhD 21, Dr David A Simpson PhD 6, and Dr Chris Williams MB BS33.

<u>Samples, logistics, leadership and supervision:</u>

Dr Cressida Auckland FRCPath 50, Dr John Boyes MBChB83, Dr Samir Dervisevic FRCPath52, Professor Sian Ellard FRCPath49, 50, Dr Sonia Goncalves1, Dr Emma J Meader FRCPath 51, Dr Peter Muir PhD2, Dr Husam Osman PhD 95, Reenesh Prakash MPH52, Dr Venkat Sivaprakasam PhD18, and Dr Ian B Vipond PhD2.

<u>Leadership, supervision and visualisation</u>

Dr Jane AH Masoli MBChB 49, 50.

<u>Sequencing, analysis and metadata curation</u>

Dr Nabil-Fareed Alikhan PhD 51, Matthew Carlile BSc 54, Dr Noel Craine DPhil 33, Dr Sam T Haldenby PhD 46, Dr Nadine Holmes PhD 54, Professor Ronan A Lyons MD 37, Dr Christopher Moore PhD 54, Malorie Perry MSc 33, Dr Ben Warne MRCP80, and Dr Thomas Williams MD 19.

<u>Samples, logistics and metadata curation:</u>

Dr Lisa Berry PhD 72, Dr Andrew Bosworth PhD 95, Dr Julianne Rose Brown PhD40, Sharon Campbell MSc 67, Dr Anna Casey PhD 17, Dr Gemma Clark PhD 56, Jennifer Collins BSc 66, Dr Alison Cox PhD 43, 44, Thomas Davis MSc 84, Gary Eltringham BSc 66, Dr Cariad Evans 38, 39, Dr Clive Graham MD 64, Dr Fenella Halstead PhD 18, Dr Kathryn Ann Harris PhD 40, Dr Christopher Holmes PhD 58, Stephanie Hutchings 2, Professor Miren Iturriza-Gomara PhD 46, Dr Kate Johnson 38, 39, Katie Jones MSc 72, Dr Alexander J Keeley MRCP 38, Dr Bridget A Knight PhD 49, 50, Cherian Koshy MSc, CSci, FIBMS 90, Steven Liggett 63, Hannah Lowe MSc 81, Dr Anita O Lucaci PhD 46, Dr Jessica Lynch PhD MBChB 25, 29, Dr Patrick C McClure PhD 55, Dr Nathan Moore MBChB 31, Matilde Mori BSc 25, 29, 32, Dr David G Partridge FRCP, FRCPath 38, 39, Pinglawathee Madona 43, 44, Hannah M Pymont MSc 2, Dr Paul Anthony Randell MBBCh 43, 44, Dr Mohammad Raza 38, 39, Felicity Ryan MSc 81, Dr Robert Shaw FRCPath 28, Dr Tim J Sloan PhD 57, and Emma Swindells BSc 65.

<u>Sequencing, analysis, Samples and logistics:</u>

Alexander Adams BSc 33, Dr Hibo Asad PhD 33, Alec Birchley MSc 33, Tony Thomas Brooks BSc (Hons) 41, Dr Giselda Bucca PhD 93, Ethan Butcher 70, Dr Sarah L Caddy PhD 13, Dr Laura G Caller PhD 2, 3, 12, Yasmin Chaudhry BSc 11, Jason Coombes BSc (HONS) 33, Michelle Cronin 33, Patricia L Dyal MPhil 41, Johnathan M Evans MSc 33,Laia Fina 33, Bree Gatica-Wilcox MPhil 33, Dr Iliana Georgana PhD 11, Lauren Gilbert A-Levels 33, Lee Graham BSc 33, Danielle C Groves BA 38, Grant Hall BSc 11, Ember Hilvers MPH33, Dr Myra Hosmillo PhD 11, Hannah Jones 33, Sophie Jones MSc 33, Fahad A Khokhar BSc 13, Sara Kumziene-Summerhayes MSc 33, George MacIntyre-Cockett BSc 26, Dr Rocio T Martinez Nunez PhD 94, Dr Caoimhe McKerr PhD 33, Dr Claire McMurray PhD 15, Dr Richard Myers 7, Yasmin Nicole Panchbhaya BSc 41, Malte L Pinckert MPhil 11, Amy Plimmer 33, Dr Joanne Stockton PhD 15, Sarah Taylor 33, Dr Alicia Thornton 7, Amy Trebes MSc 26, Alexander J Trotter MRes 51, Helena Jane Tutill BSc 41, Charlotte A Williams BSc 41, Anna Yakovleva BSc 11 and Dr Wen C Yew PhD 62.

<u>Sequencing, analysis and software and analysis tools:</u>

Dr Mohammad T Alam PhD 71, Dr Laura Baxter PhD 71, Olivia Boyd MSc 96, Dr Fabricia F. Nascimento PhD 96, Timothy M Freeman MPhil 38, Lily Geidelberg MSc 96, Dr Joseph Hughes PhD 21, David Jorgensen MSc 96, Dr Benjamin B Lindsey MRCP 38, Dr Richard J Orton PhD 21, Dr Manon Ragonnet-Cronin PhD 96 Joel Southgate MSc 33, 34, and Dr Sreenu Vattipally PhD 21.

<u>Samples, logistics and software and analysis tools:</u>

Dr Igor Starinskij MSc MRCP 23.

<u>Visualisation and software and analysis tools:</u>

Dr Joshua B Singer PhD 21, Dr Khalil Abudahab PhD 1, 30, Leonardo de Oliveira Martins PhD51, Dr Thanh Le-Viet PhD 51, Mirko Menegazzo 30, Ben EW Taylor Meng 1, 30, and Dr Corin A Yeats PhD 30.

<u>Project Administration:</u>

Sophie Palmer 3, Carol M Churcher 3, Dr Alisha Davies 33, Elen De Lacy MSc 33, Fatima Downing 33, Sue Edwards 33, Dr Nikki Smith PhD 38, Dr Francesc Coll PhD97, Dr Nazreen F Hadjirin PhD3 and Dr Frances Bolt PhD 44, 45.

<u>Leadership and supervision:</u>

Dr. Alex Alderton 1, Dr Matt Berriman 1, Ian G Charles 51, Dr Nicholas Cortes MBChB 31, Dr Tanya Curran PhD 88, Prof John Danesh 1, Dr Sahar Eldirdiri MBBS, MSC FRCPath 84, Dr Ngozi Elumogo FRCPath 52, Prof Andrew Hattersley FRS 49, 50, Professor Alison Holmes MD 44, 45, Dr Robin Howe 33, Dr Rachel Jones 33, Anita Kenyon MSc 84, Prof Robert A Kingsley PhD 51, Professor Dominic Kwiatkowski 1, 9, Dr Cordelia Langford1, Dr Jenifer Mason MBBS 48, Dr Alison E Mather PhD 51, Lizzie Meadows MA 51, Dr Sian Morgan FRCPath 36, Dr James Price PhD 44, 45, Trevor I Robinson MSc 48 * Dr Giri Shankar 33, John Wain 51, and Dr Mark A Webber PhD51.

<u>Metadata curation:</u>

Dr Declan T Bradley PhD 5, 6, Dr Michael R Chapman PhD 1, 3, 4, Dr Derrick Crooke 28, Dr David Eyre PhD 28, Professor Martyn Guest PhD34, Huw Gulliver 34, Dr Sarah Hoosdally 28, Dr Christine Kitchen PhD 34, Dr Ian Merrick PhD 34, Siddharth Mookerjee MPH 44, 45, Robert Munn BSc 34, Professor Timothy Peto PhD28, Will Potter 52, Dr Dheeraj K Sethi MBBS 52, Wendy Smith 56, Dr Luke B Snell MB BS 75, 94, Dr Rachael Stanley PhD 52, Claire Stuart 52 and Dr Elizabeth Wastenge MD 20.

<u>Sequencing and analysis:</u>

Dr Erwan Acheson PhD 6, Safiah Afifi BSc 36, Dr Elias Allara MD PhD 2, 3, Dr Roberto Amato 1, Dr Adrienn Angyal PhD38, Dr Elihu Aranday-Cortes PhD/DVM21, Cristina Ariani 1, Jordan Ashworth 19, Dr Stephen Attwood 24, Alp Aydin MSci 51, David J Baker BEng 51, Dr Carlos E Balcazar PhD 19, Angela Beckett MSc 68 Robert Beer BSc 36, Dr Gilberto Betancor PhD76, Emma Betteridge 1, Dr David Bibby 7, Dr Daniel Bradshaw 7, Catherine Bresner Bsc(Hons) 34, Dr Hannah E Bridgewater PhD 71, Alice Broos BSc (Hons) 21, Dr Rebecca Brown PhD 38, Dr Paul E Brown PhD 71, Dr Kirstyn Brunker PhD 22, Dr Stephen N Carmichael PhD 21, Jeffrey K. J. Cheng MSc 71, Dr Rachel Colquhoun DPhil 19, Dr Gavin Dabrera 7, Dr Johnny Debebe PhD 54, Eleanor Drury 1, Dr Louis du Plessis 24, Richard Eccles MSc 46, Dr Nicholas Ellaby 7, Audrey Farbos MSc 49, Ben Farr 1, Dr Jacqueline Findlay PhD 41, Chloe L Fisher MSc 74, Leysa Marie Forrest MSc 41, Dr Sarah Francois 24, Lucy R. Frost BSc 71, William Fuller BSc 34, Dr Eileen Gallagher 7, Dr Michael D Gallagher PhD19, Matthew Gemmell MSc 46, Dr Rachel AJ Gilroy PhD 51, Scott Goodwin 1, Dr Luke R Green PhD 38, Dr Richard Gregory PhD 46, Dr Natalie Groves 7, Dr James W Harrison PhD 49, Hassan Hartman 7, Dr Andrew R Hesketh PhD93,Verity Hill 19, Dr Jonathan Hubb 7, Dr Margaret

Hughes PhD46, Dr David K Jackson 1, Dr Ben Jackson PhD 19, Dr Keith James 1, Natasha Johnson BSc (Hons)21, Ian Johnston 1, Jon-Paul Keatley 1, Dr Moritz Kraemer 24, Dr Angie Lackenby 7, Dr Mara Lawniczak 1, Dr David Lee 7, Rich Livett 1, Stephanie Lo 1, Daniel Mair BSc (Hons) 21, Joshua Maksimovic FD sport science 36, Nikos Manesis 7, Dr Robin Manley Ph.D 49, Dr Carmen Manso 7, Dr Angela Marchbank BSc 34, Dr Inigo Martincorena 1, Dr Tamyo Mbisa 7, Kathryn McCluggage MSC 36,Dr JT McCrone PhD 19, Shahjahan Miah 7, Michelle L Michelsen BSc 49, Dr Mari Morgan PhD 33, Dr Gaia Nebbia PhD, FRCPath 78,Charlotte Nelson MSc 46, Jenna Nichols BSc (Hons) 21, Dr Paola Niola PhD 41, Dr Kyriaki Nomikou PhD21, Steve Palmer 1, Dr. Naomi Park 1, Dr Yasmin A Parr PhD21, Dr Paul J Parsons PhD 38, Vineet Patel 7, Dr. Minal Patel 1, Clare Pearson MSc 2, 1, Dr Steven Platt 7, Christoph Puethe 1, Dr. Mike Quail 1,Dr JaynaRaghwani 24, Dr Lucille Rainbow PhD 46, Shavanthi Rajatileka 1, Dr Mary Ramsay 7, Dr Paola C Resende Silva PhD 41, 42, Steven Rudder 51, Dr Chris Ruis 3, Dr Christine M Sambles PhD 49, Dr Fei Sang PhD 54, Dr Ulf Schaefer7, Dr Emily Scher PhD 19, Dr. Carol Scott 1, Lesley Shirley 1, Adrian W Signell BSc 76, John Sillitoe 1, Christen Smith 1, Dr Katherine L Smollett PhD 21, Karla Spellman FD 36, Thomas D Stanton BSc 19, Dr David J Studholme PhD 49, Ms Grace Taylor-Joyce BSc 71, Dr Ana P Tedim PhD 51, Dr Thomas Thompson PhD6, Dr Nicholas M Thomson PhD 51, Scott Thurston1, Lily Tong PhD 21, Gerry Tonkin-Hill 1, Rachel M Tucker MSc 38, Dr Edith E Vamos PhD 4,Dr Tetyana Vasylyeva24, Joanna Warwick-Dugdale BSc 49, Danni Weldon 1, Dr Mark Whitehead PhD 46, Dr David Williams 7,Dr Kathleen A Williamson PhD19,Harry D Wilson BSc 76,Trudy Workman HNC 34, Dr Muhammad Yasir PhD 51, Dr Xiaoyu Yu PhD 19, and Dr Alex Zarebski 24.

<u>Samples and logistics:</u>

Dr Evelien M Adriaenssens PhD 51, Dr Shazaad S Y Ahmad MSc 2, 47, Adela Alcolea-Medina MPharm 59, 77, Dr John Allan PhD60, Dr Patawee Asamaphan PhD21, Laura Atkinson MSc 40, Paul Baker MD 63, Professor Jonathan Ball PhD 55, Dr Edward Barton MD64, Dr. Mathew A Beale1, Dr. Charlotte Beaver1 * Dr Andrew Beggs PhD16, Dr Andrew Bell PhD51, Duncan J Berger 1, Dr Louise Berry. 56, Claire M Bewshea MSc 49, Kelly Bicknell 70, Paul Bird 58, Dr Chloe Bishop 7, Dr Tim Boswell 56, Cassie Breen BSc48, Dr Sarah K Buddenborg1, Dr Shirelle Burton-Fanning MD 66, Dr Vicki Chalker 7, Dr Joseph G Chappell PhD 55, Themoula Charalampous MSc 78, 94, Claire Cormie3, Dr Nick Cortes PhD29, 25, Dr Lindsay J Coupland PhD 52, Angela Cowell MSc48, Dr Rose K Davidson PhD 53, Joana Dias MSc3, Dr Maria Diaz PhD51, Thomas Dibling1, Matthew J Dorman1, Dr Nichola Duckworth57, Scott Elliott70, Sarah Essex63, Karlie Fallon 58, Theresa Feltwell 8, Dr Vicki M Fleming PhD 56, Sally Forrest BSc 3, Luke Foulser1, Maria V Garcia-Casado1, Dr Artemis Gavriil PhD 41, Dr Ryan P George PhD47, Laura Gifford MSc 33, Harmeet K Gill PhD3, Jane Greenaway MSc65, Luke Griffith Bsc53, Ana Victoria Gutierrez51, Dr Antony D Hale MBBS85, Dr Tanzina Haque FRCPath, PhD91, Katherine L Harper MBiol85, Dr Ian Harrison 7, Dr Judith Heaney PhD89, Thomas Helmer 58, Ellen E Higginson PhD 3, Richard Hopes 2, Dr Hannah C Howson-Wells PhD 56, Dr Adam D Hunter 1, Robert Impey 70, Dr Dianne Irish-Tavares FRCPath 91, David A Jackson1, Kathryn A Jackson MSc 46, Dr Amelia Joseph 56, Leanne Kane 1, Sally Kay 1, Leanne M Kermack MSc 3, Manjinder Khakh 56, Dr Stephen P Kidd PhD29, 25,31,, Dr Anastasia Kolyva PhD 51, Jack CD Lee BSc 40, Laura Letchford 1, Nick Levene MSc79, Dr LisaJ Levett PhD 89, Dr Michelle M Lister PhD 56, Allyson Lloyd 70, Dr Joshua Loh PhD60, Dr Louissa R Macfarlane-Smith PhD85, Dr Nicholas W Machin MSc 2, 47, Mailis Maes M.phil3, Dr Samantha McGuigan 1, Liz McMinn 1, Dr Lamia Mestek-Boukhibar D.Phil 41, Dr Zoltan Molnar PhD 6, Lynn Monaghan 79, Dr Catrin Moore 27, Plamena Naydenova BSc 3, Alexandra S Neaverson 1, Dr. Rachel Nelson PhD 1, Marc O Niebel MSc21, Elaine O’Toole BSc 48, Debra Padgett BSc 64, Gaurang Patel 1, Dr Brendan AI Payne MD 66, Liam Prestwood 1, Dr Veena Raviprakash MD67, Nicola Reynolds PhD86 Dr Alex Richter PhD 16, Dr Esther Robinson PhD95, Dr Hazel A Rogers1, Dr Aileen Rowan PhD 96, Garren Scott BSc 64, Dr Divya Shah PhD40, Nicola Sheriff BSc 67, Dr Graciela Sluga MD - MSc92, Emily Souster1, Dr. Michael Spencer-Chapman1, Sushmita Sridhar BSc1, 3, Tracey Swingler 53, Dr Julian Tang58, Professor Graham P Taylor DSc96, Dr Theocharis Tsoleridis PhD55, Dr Lance Turtle PhD MRCP46, Dr Sarah Walsh 57, Dr Michelle Wantoch PhD 86, Joanne Watts BSc48, Dr Sheila Waugh MD66, Sam Weeks41, Dr Rebecca Williams BMBS 31, Dr Iona Willingham56, Dr Emma L Wise PhD 25, 29, 31, Victoria Wright BSc 54, Dr Sarah Wyllie 70, and Jamie Young BSc 3.

<u>Software and analysis tools</u>

Amy Gaskin MSc33, Dr Will Rowe PhD 15, and Dr Igor Siveroni PhD96.

<u>Visualisation:</u>

Dr Robert Johnson PhD 96.

1 Wellcome Sanger Institute, 2 Public Health England, 3 University of Cambridge, 4 Health Data Research UK, Cambridge, 5 Public Health Agency, Northern Ireland, 6 Queen’s University Belfast 7 Public Health England Colindale, 8 Department of Medicine, University of Cambridge, 9 University of Oxford, 10 Departments of Infectious Diseases and Microbiology, Cambridge University Hospitals NHS Foundation Trust; Cambridge, UK, 11 Division of Virology, Department of Pathology, University of Cambridge, 12 The Francis Crick Institute, 13 Cambridge Institute for Therapeutic Immunology and Infectious Disease, Department of Medicine, 14 Public Health England, Clinical Microbiology and Public Health Laboratory, Cambridge, UK, 15 Institute of Microbiology and Infection, University of Birmingham, 16 University of Birmingham, 17 Queen Elizabeth Hospital, 18 Heartlands Hospital, 19 University of Edinburgh, 20 NHS Lothian, 21 MRC-University of Glasgow Centre for Virus Research, 22 Institute of Biodiversity, Animal Health & Comparative Medicine, University of Glasgow, 23 West of Scotland Specialist Virology Centre, 24 Dept Zoology, University of Oxford, 25 University of Surrey, 26 Wellcome Centre for Human Genetics, Nuffield Department of Medicine, University of Oxford, 27 Big Data Institute, Nuffield Department of Medicine, University of Oxford, 28 Oxford University Hospitals NHS Foundation Trust, 29 Basingstoke Hospital, 30 Centre for Genomic Pathogen Surveillance, University of Oxford, 31 Hampshire Hospitals NHS Foundation Trust, 32 University of Southampton, 33 Public Health Wales NHS Trust, 34 Cardiff University, 35 Betsi Cadwaladr University Health Board, 36 Cardiff and Vale University Health Board, 37 Swansea University, 38 University of Sheffield, 39 Sheffield Teaching Hospitals, 40 Great Ormond Street NHS Foundation Trust, 41 University College London, 42 Oswaldo Cruz Institute, Rio de Janeiro 43 North West London Pathology, 44 Imperial College Healthcare NHS Trust, 45 NIHR Health Protection Research Unit in HCAI and AMR, Imperial College London, 46 University of Liverpool, 47 Manchester University NHS Foundation Trust, 48 Liverpool Clinical Laboratories, 49 University of Exeter, 50 Royal Devon and Exeter NHS Foundation Trust, 51 Quadram Institute Bioscience, University of East Anglia, 52 Norfolk and Norwich University Hospital, 53 University of East Anglia, 54 Deep Seq, School of Life Sciences, Queens Medical Centre, University of Nottingham, 55 Virology, School of Life Sciences, Queens Medical Centre, University of Nottingham, 56 Clinical Microbiology Department, Queens Medical Centre, 57 PathLinks, Northern Lincolnshire & Goole NHS Foundation Trust, 58 Clinical Microbiology, University Hospitals of Leicester NHS Trust, 59 Viapath, 60 Hub for Biotechnology in the Built Environment, Northumbria University, 61 NU-OMICS Northumbria University, 62 Northumbria University, 63 South Tees Hospitals NHS Foundation Trust, 64 North Cumbria Integrated Care NHS Foundation Trust, 65 North Tees and Hartlepool NHS Foundation Trust, 66 Newcastle Hospitals NHS Foundation Trust, 67 County Durham and Darlington NHS Foundation Trust, 68 Centre for Enzyme Innovation, University of Portsmouth, 69 School of Biological Sciences, University of Portsmouth, 70 Portsmouth Hospitals NHS Trust, 71 University of Warwick, 72 University Hospitals Coventry and Warwickshire, 73 Warwick Medical School and Institute of Precision Diagnostics, Pathology, UHCW NHS Trust, 74 Genomics Innovation Unit, Guy’s and St. Thomas’ NHS Foundation Trust, 75 Centre for Clinical Infection & Diagnostics Research, St. Thomas’ Hospital and Kings College London, 76 Department of Infectious Diseases, King’s College London, 77 Guy’s and St. Thomas’ Hospitals NHS Foundation Trust, 78 Centre for Clinical Infection and Diagnostics Research, Department of Infectious Diseases, Guy’s and St Thomas’ NHS Foundation Trust, 79 Princess Alexandra Hospital Microbiology Dept., 80 Cambridge University Hospitals NHS Foundation Trust, 81 East Kent Hospitals University NHS Foundation Trust, 82 University of Kent, 83 Gloucestershire Hospitals NHS Foundation Trust, 84 Department of Microbiology, Kettering General Hospital, 85 National Infection Service, PHE and Leeds Teaching Hospitals Trust, 86 Cambridge Stem Cell Institute, University of Cambridge, 87 Public Health Scotland, 88 Belfast Health & Social Care Trust, 89 Health Services Laboratories, 90 Barking, Havering and Redbridge University Hospitals NHS Trust, 91 Royal Free NHS Trust, 92 Maidstone and Tunbridge Wells NHS Trust, 93 University of Brighton, 94 Kings College London, 95 PHE Heartlands, 96 Imperial College London, 97 Department of Infection Biology, London School of Hygiene and Tropical Medicine.

