## Supplementary figures and images for "Combined epidemiological and genomic analysis of nosocomial SARS-CoV-2 transmission identifies community social distancing as the dominant intervention reducing outbreaks"

### Supplemental Figure S1

#### Supplemental Figure 1

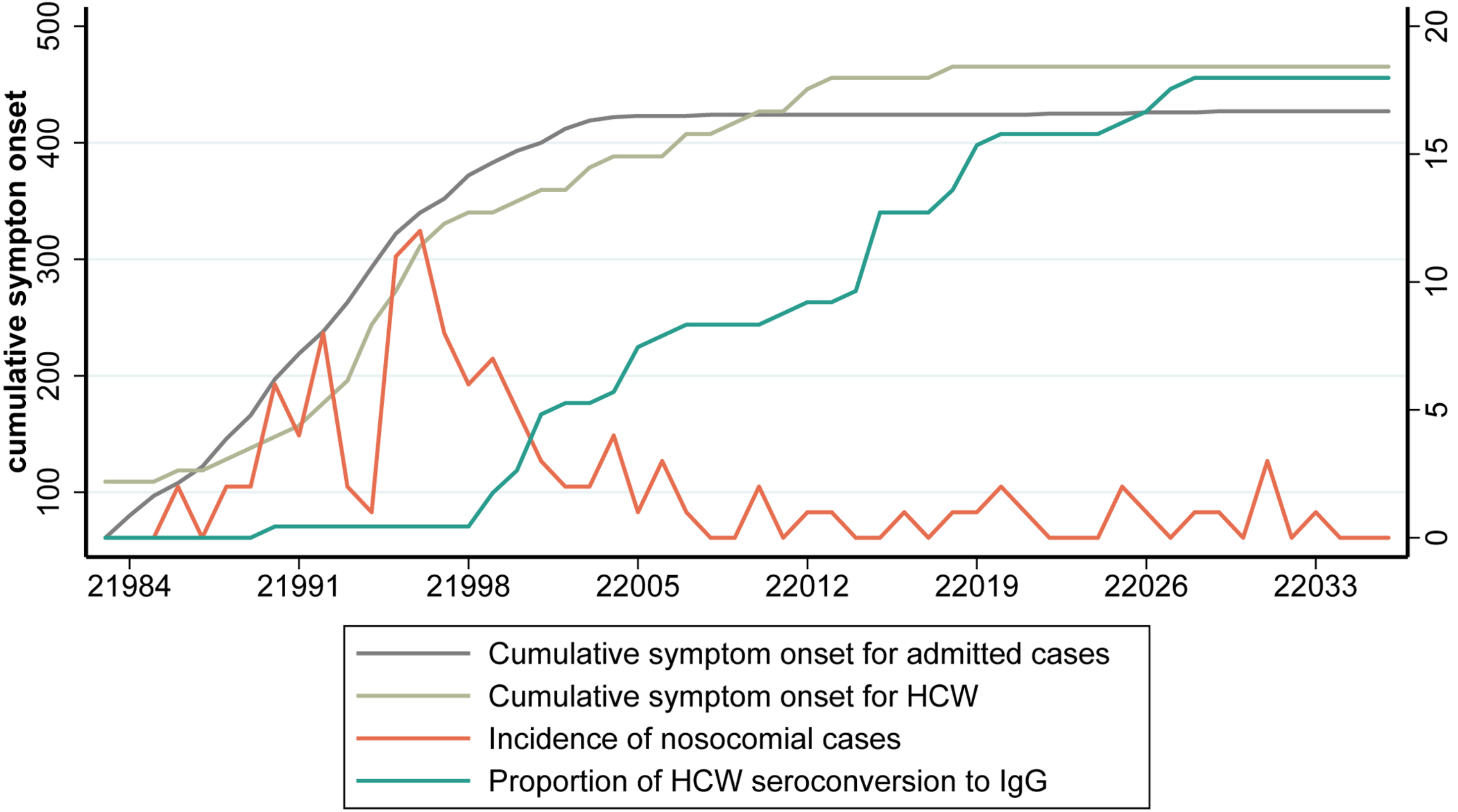

### Supplemental Figure S3

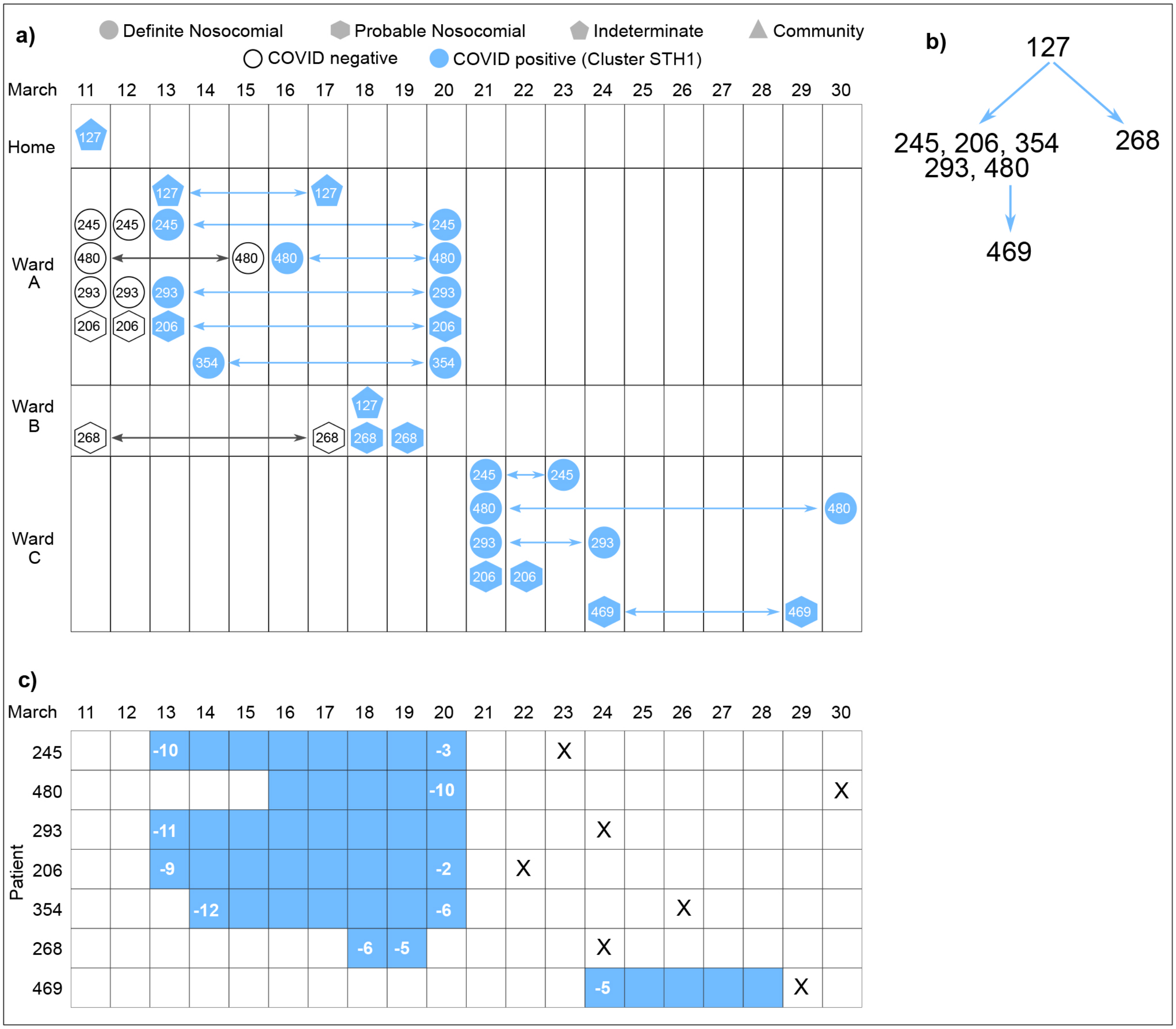

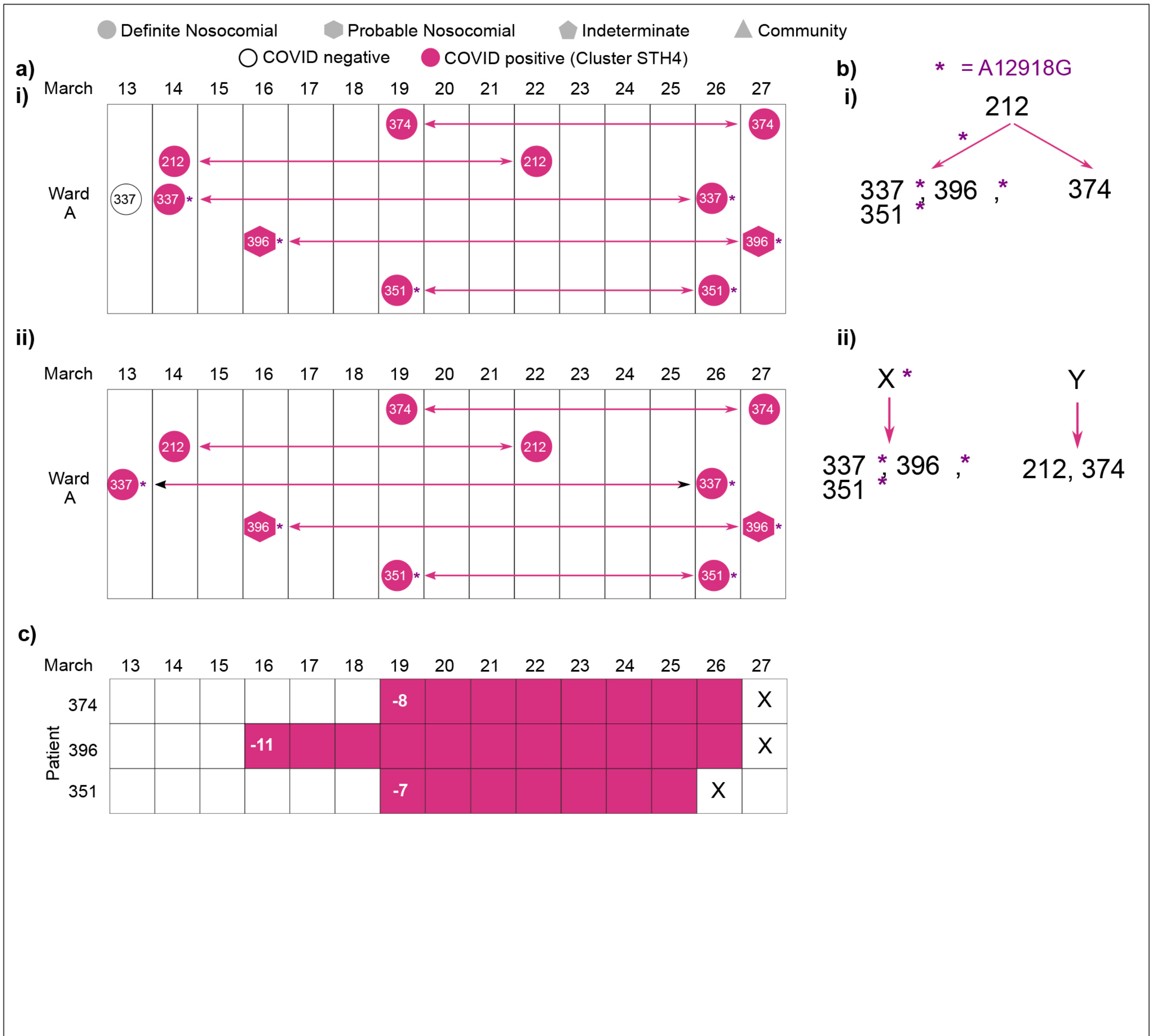

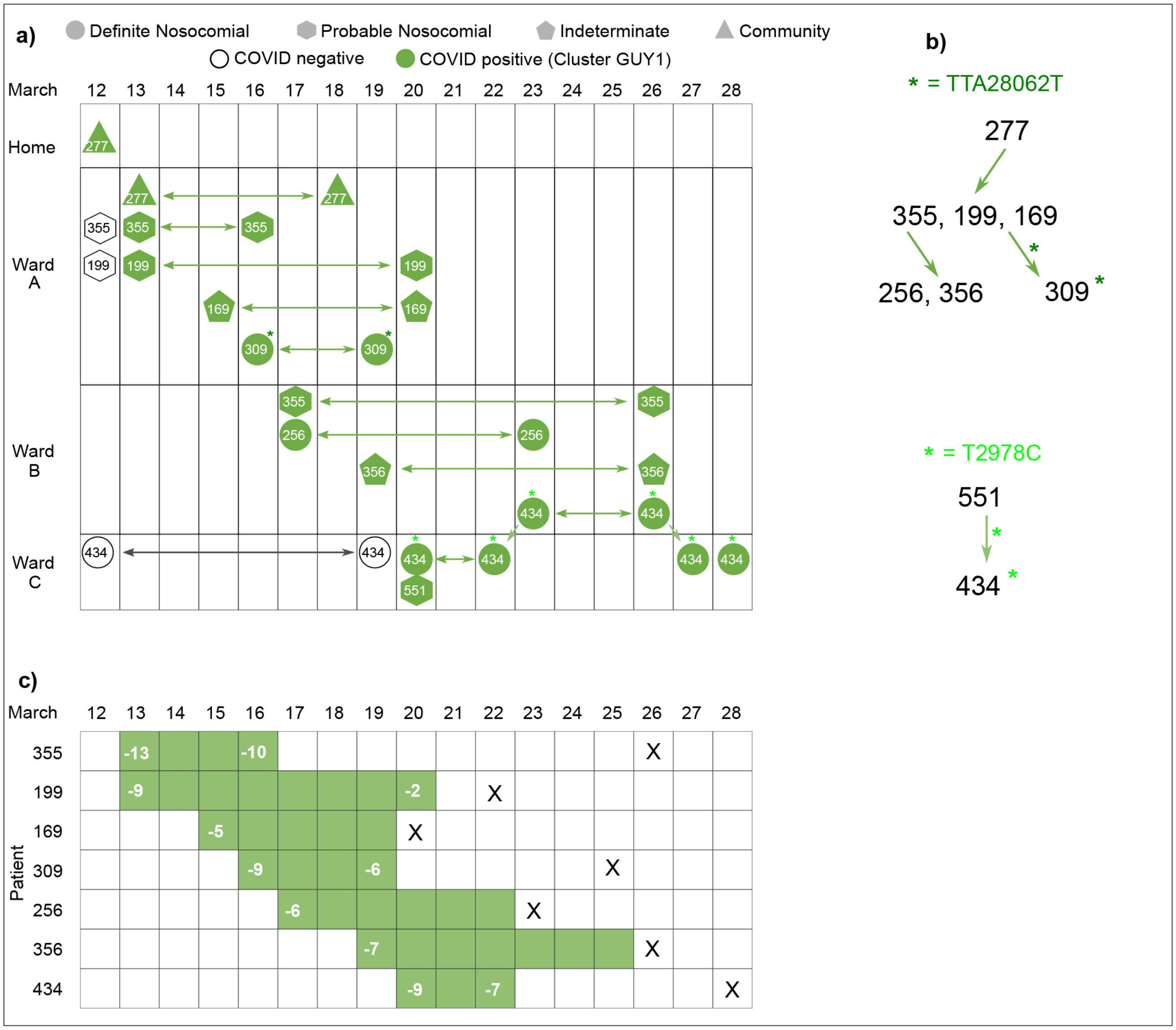


| March | 12 | 13 | 14 | 15 | 16 | 17 | 18 | 19 | 20 | 21 | 22 | 23 | 24 | 25 | 26 | 27 | 28 |
|---|---|---|---|---|---|---|---|---|---|---|---|---|---|---|---|---|---|
| 355 | | -13 | | | -10 | | | | | | | | | | X | | |
| 199 | | -9 | | | | | | | -2 | | X | | | | | | |
| 169 | | | | -5 | | | | | X | | | | | | | | |
| 309 | | | | | -9 | | | -6 | | | | | | X | | | |
| 256 | | | | | | -6 | | | | | | X | | | | | |
| 356 | | | | | | | | -7 | | | | | | | X | | |
| 434 | | | | | | | | | -9 | | -7 | | | | | | X |

Figure S3C

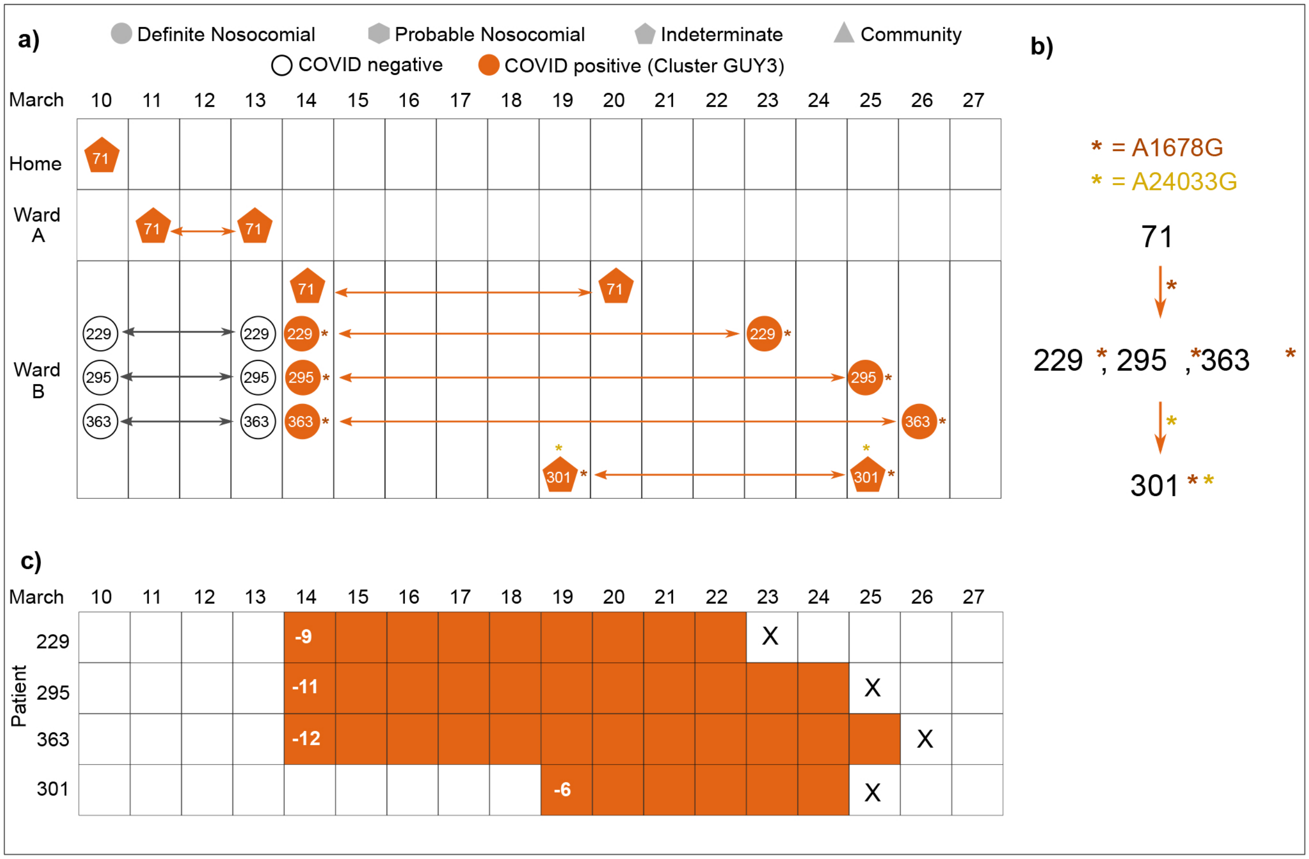
