## Supplemental Figure S2 for "Combined epidemiological and genomic analysis of nosocomial SARS-CoV-2 transmission identifies community social distancing as the dominant intervention reducing outbreaks"

SUPPLEMENTARY FIGURE 2. Cluster STH1

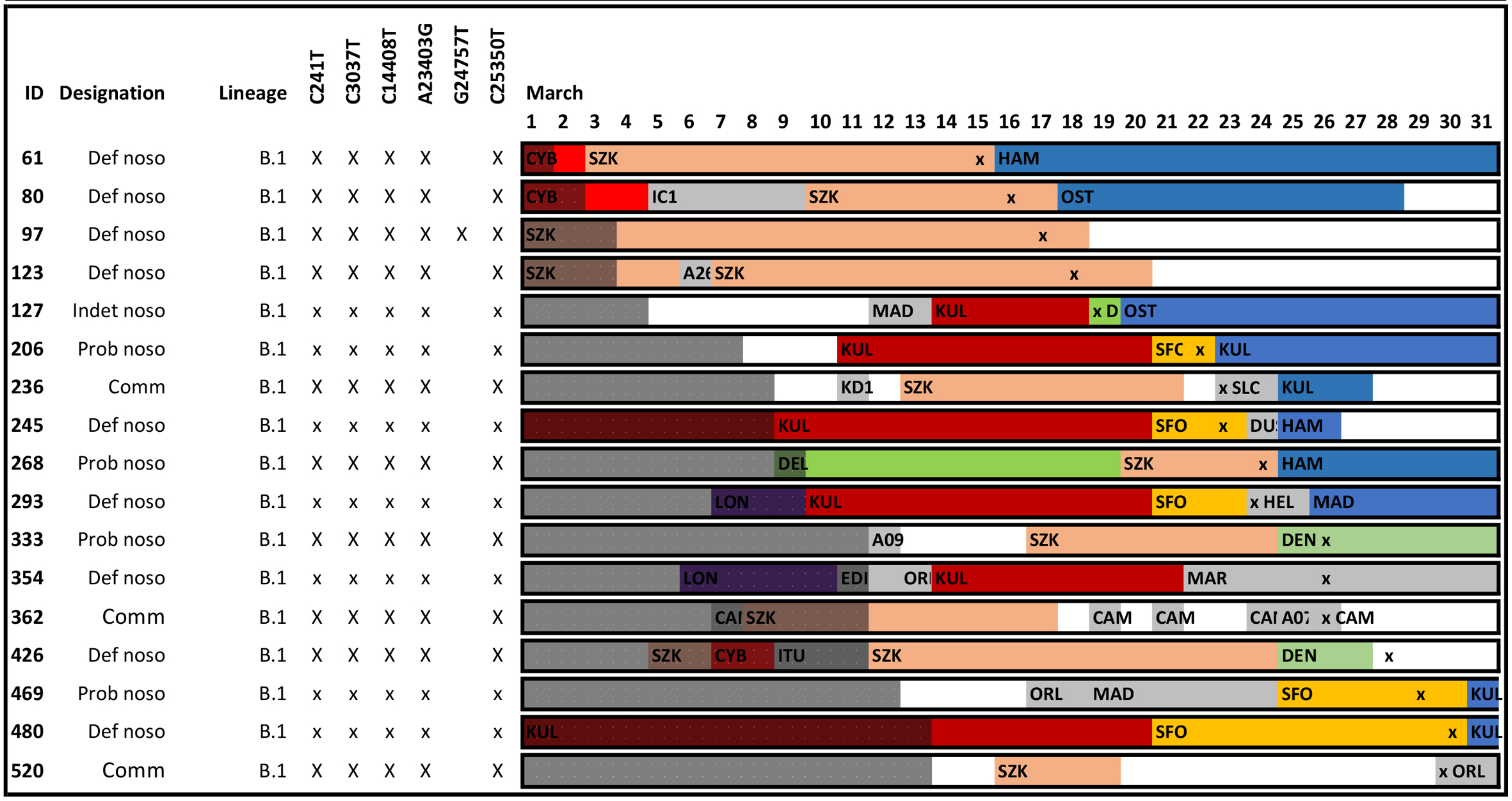


SUPPLEMENTARY FIGURE 2. Cluster STH2

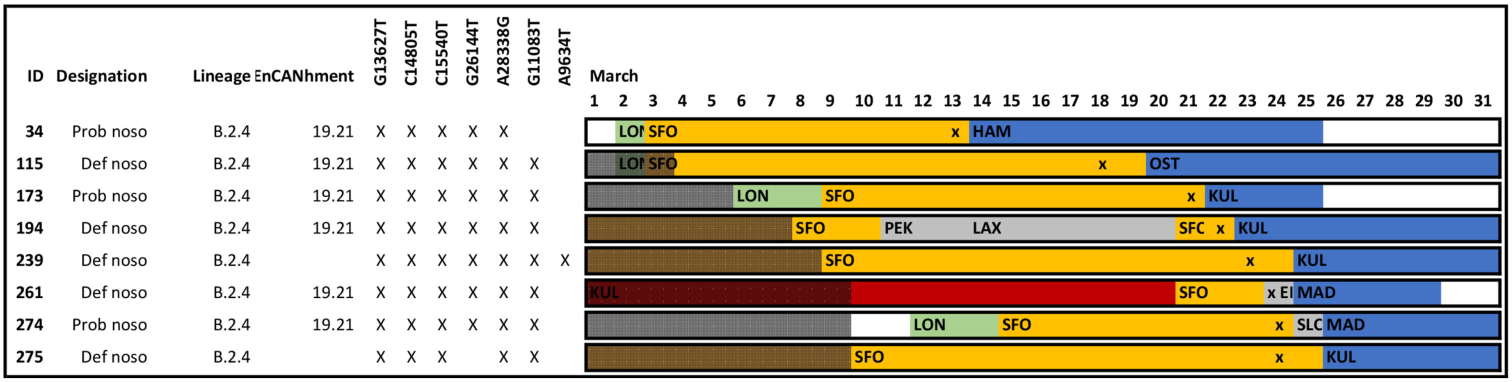


SUPPLEMENTARY FIGURE 2. Cluster STH3

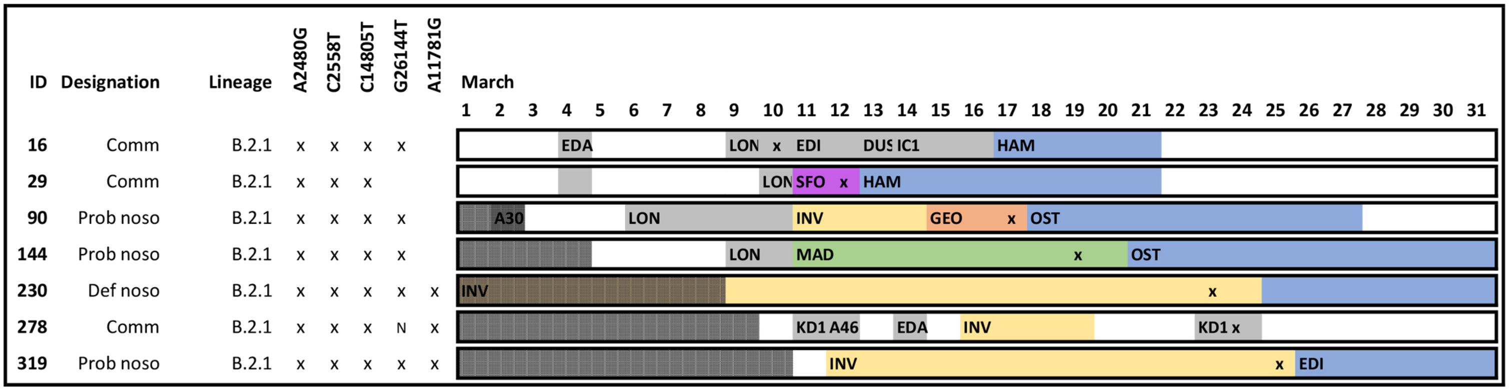

### SUPPLEMENTARY FIGURE 2. Cluster STH4

| ID | Designation | Lineage | A2480G | C2558T | C6070T | A12918G | C14805T | G26144T | G11083T | March (1–31) |
|---|---|---|---|---|---|---|---|---|---|---|
| 212 | Def noso | B.2.1 | x | x | x | | x | x | | KUL · DEN · x IC DEL · DEN · HAM |
| 337 | Def noso | B.2.1 | x | x | x | x | x | x | x | DEN · x · DFW |
| 351 | Def noso | B.2.1 | x | x | x | x | x | x | x | IC1 · DOI · DEN · x |
| 374 | Def noso | B.2.1 | x | x | x | | x | x | x | IC6 · DEL · DEN · x |
| 396 | Prob noso | B.2.1 | x | x | x | x | x | x | x | DEN · x · NOR |

### SUPPLEMENTARY FIGURE 2. Cluster STH5

| ID | Designation | Lineage | C241T | C3037T | C14408T | A23403G | G28881A | G28882A | G28883C | March (1–31) |
|---|---|---|---|---|---|---|---|---|---|---|
| 141 | Comm | B.1.1 | x | x | x | x | x | x | x | x SZK |
| 205 | Prob noso | B.1.1 | x | x | x | x | x | x | x | SZK · x D · HAM |

### SUPPLEMENTARY FIGURE 2. Cluster GUY2

| ID | Designation | Lineage | C241T | C1150T | C3037T | T2978C | C14408T | A22481T | A23403G | TTA28062T | G28881A | G28882A | G28883C | March (1–31) |
|---|---|---|---|---|---|---|---|---|---|---|---|---|---|---|
| 196 | Indet noso | B.1.1 | x | x | x | | x | x | x | | x | x | x | SAL · DOR · x HND · PEK |
| 322 | Def noso | B.1.1 | x | x | x | | x | x | x | | x | x | x | DOR · BLL · DOR · x |

### SUPPLEMENTARY FIGURE 2. Cluster GUY3

| ID | Designation | Lineage | A1678G | C3096T | G13627T | C14805T | C15540T | A24033G | G26144T | A28338G | March (1–31) |
|---|---|---|---|---|---|---|---|---|---|---|---|
| 71 | Indet noso | B.2.4 | | x | x | x | x | | x | x | IC9 · x HKG · PEK |
| 229 | Def noso | B.2.4 | x | x | x | x | x | | x | x | HKG · x · PEK |
| 295 | Def noso | B.2.4 | x | x | x | x | x | | x | x | HKG · x · PEK |
| 301 | Indet noso | B.2.4 | x | x | x | x | x | x | x | x | HKG · x · PEK |
| 363 | Def noso | B.2.4 | x | | | x | | | x | x | HKG · x · PEK |

### SUPPLEMENTARY FIGURE 2. Cluster GUY4

| ID | Designation | Lineage | A2480G | C2558T | G9188T | C14805T | G26144T | March (1–31) |
|---|---|---|---|---|---|---|---|---|
| 200 | Def noso | B.2.1 | x | x | x | x | x | HKG · x · PEK |
| 302 | Def noso | B.2.1 | x | x | | x | x | HKG · x |
| 372 | Def noso | B.2.1 | x | x | | x | x | AMS · x H · PEK |

**SUPPLEMENTARY FIGURE 2. Cluster GUY5**

| ID | Designation | Lineage | C241T | C2306T | C3037T | C3784T | C14408T | G18653A | C22000T | A23403G | G28881A | G28882A | G28883C |
|---|---|---|---|---|---|---|---|---|---|---|---|---|---|
| 471 | Def noso | B.1.1 | x | x | x | x | x | x | | x | x | x | x |
| 530 | Def noso | B.1.1 | x | x | x | x | x | x | x | x | x | x | x |
| 559 | Prob noso | B.1.1 | x | x | x | x | x | x | | x | x | x | x |

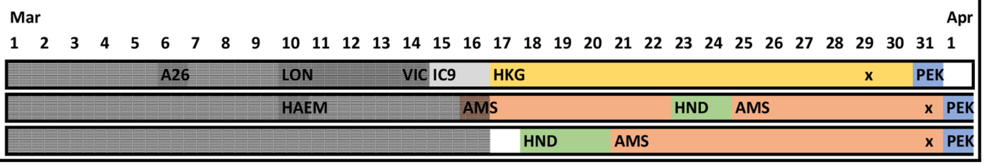


SUPPLEMENTARY FIGURE 2. Cluster GUY6

| ID | Designation | Lineage | C241T | C3037T | C14408T | A23403G | G25429T |
|---|---|---|---|---|---|---|---|
| 367 | Def noso | B.1 | x | x | x | x | x |
| 557 | Prob noso | B.1 | x | x | x | x | x |

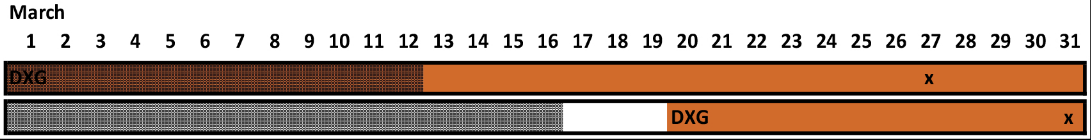


**SUPPLEMENTARY FIGURE 2. Cluster GUY7**

| ID | Designation | Lineage | T1490A | C14805T | T17247C | G25483T | G26144T | T26870G |
|---|---|---|---|---|---|---|---|---|
| 235 | Def noso | B.1.1 | x | x | x | x | x | x |
| 255 | Def noso | B.1.1 | x | x | x | | x | x |

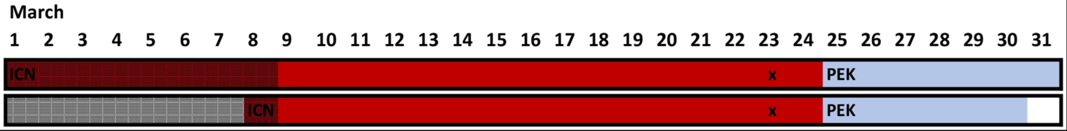


**SUPPLEMENTARY FIGURE 2. Cluster INB**

| ID | Designation | Lineage | C1593T | A2480G | C2558T | C14805T | G26144T |
|---|---|---|---|---|---|---|---|
| 94 | Def noso | B.2.1 | | x | x | x | x |
| 150 | Def noso | B.2.1 | x | x | x | x | x |
| 153 | Def noso | B.2.1 | | x | x | x | N |
| 154 | Def noso | B.2.1 | | x | x | x | x |
| 155 | Def noso | B.2.1 | | x | x | x | x |
| 216 | Def noso | B.2.1 | | x | x | x | ? |
| 217 | Def noso | B.2.1 | | x | x | x | x |

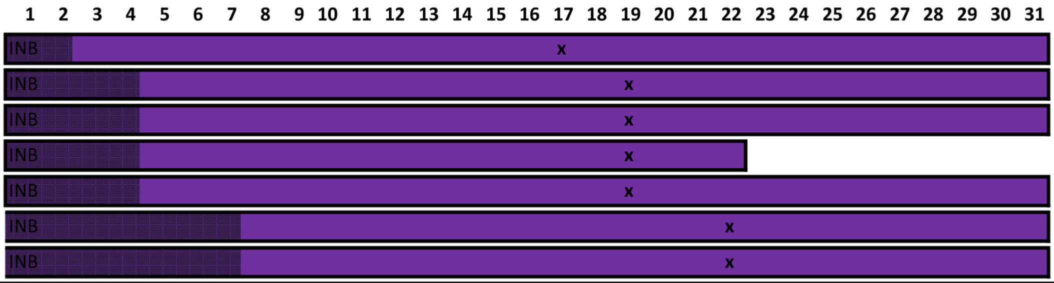


**SUPPLEMENTARY FIGURE 2. Cluster IND**

| ID | Designation | Lineage | A2480G | C2558T | C14805T | G22094A | A22252G | G26144T |
|---|---|---|---|---|---|---|---|---|
| 195 | Def noso | B.2.1 | x | x | x | x | x | |
| 279 | Def noso | B.2.1 | x | x | x | x | x | |
| 282 | Prob noso | B.2.1 | x | x | x | x | x | x |

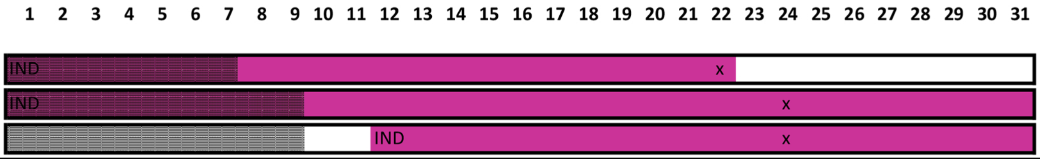
